# Emergence of immune escape at dominant SARS-CoV-2 killer T-cell epitope

**DOI:** 10.1101/2021.06.21.21259010

**Authors:** Garry Dolton, Cristina Rius, Md Samiul Hasan, Aaron Wall, Barbara Szomolay, Enas Behiry, Thomas Whalley, Joel Southgate, Anna Fuller, The COVID-19 Genomics UK (COG-UK) consortium, Théo Morin, Katie Topley, Li Rong Tan, Philip J. R. Goulder, Owen B. Spiller, Pierre J. Rizkallah, Lucy C. Jones, Thomas R. Connor, Andrew K. Sewell

## Abstract

The adaptive immune system protects against infection via selection of specific antigen receptors on B-cells and T-cells. We studied the prevalent CD8 ‘killer’ T-cell response mounted against SARS-CoV-2 Spike_269-277_ epitope YLQPRTFLL via the most frequent Human Leukocyte Antigen (HLA) class I worldwide, HLA A*02. The widespread Spike P272L mutation has arisen in at least 14 different SARS-CoV-2 lineages to date, including in lineages identified as variants of concern. P272L was common in the B.1.177 lineage associated with establishing the ‘second wave’ in Europe. The large CD8 T-cell response seen across a cohort of HLA A*02^+^ convalescent patients, comprising of over 120 different TCRs, failed to respond to the P272L. Sizable populations (0.01%-0.2%) of total CD8 T-cells from individuals vaccinated against SARS-CoV-2 stained with HLA A*02-YLQPRTFLL multimers but failed to bind to the P272L reagent. Viral escape at prevalent T-cell epitopes restricted by high frequency HLA may be particularly problematic when vaccine immunity is focussed on a single protein such as SARS-CoV-2 Spike and provides a strong argument for inclusion of multiple viral proteins in next generation vaccines and highlights the urgent need for monitoring T-cell escape in new SARS-CoV-2 variants.

**New in Version 2:**

- Updated as P272L now seen in 14 different SARS-CoV-2 lineages including the B.1.1.7/Alpha (UK variant) lineage
- Atomic structures of HLA A*02-YLQPRTFLL and HLA A*02-YLQ**<u>L</u>**RTFLL added
- Pierre Rizkallah added as an author and author list changed to reflect the contributions of Aaron Wall and Anna Fuller to the newly added datasets

## INTRODUCTION

The mammalian immune system utilises numerous highly developed mechanisms to defend against viral infection. The most sophisticated of these systems, adaptive immunity, is controlled via vast fleets of highly variable antigen-binding molecules called B-cell receptors (antibodies) or T-cell receptors (TCRs). The advent of <u>s</u>evere <u>a</u>cute respiratory <u>s</u>yndrome <u>co</u>rona<u>v</u>irus <u>2</u> (SARS-CoV-2) as a novel human coronavirus and the stress on global healthcare systems caused by the associated <u>co</u>rona<u>v</u>irus <u>d</u>isease 2019 (COVID-19) has focussed the world’s attention on ways to combat this emerging infection. The mechanisms used by coronaviruses to escape from the host adaptive immune system are not well understood but are now firmly in the spotlight. The developing picture shows that neutralising antibodies, CD4 ‘helper’ T-cells and CD8 ‘killer’ T-cells all contribute to the control of SARS-CoV-2 and the protection offered by currently approved vaccines^1,2^. Although research focused on antibody-mediated immunity has made up the majority of published SARS-CoV-2 immunological work to date, some evidence suggests that antibodies may play a secondary role in ultimately clearing SARS-CoV-2 infection compared to T-cells. The presence of SARS-CoV-2-specific CD4 and CD8 T-cells has been reported to correlate with reduced COVID-19 severity while neutralising antibodies in the same individuals did not^2^. Two SARS-CoV-2 positive agammaglobulinemia patients who developed COVID-19 symptoms during the first wave of infection in Italy, required neither oxygen nor intensive care before making full recoveries^3^. The authors conclude that while an antibody-mediated response to SARS-CoV-2 might be important, it is not obligatory for overcoming disease^3^. This finding is consistent with multiple patients that developed COVID-19 while on B-cell depleting therapy across several studies, who resolved infection without the need for intensive treatment^4-6^. There are also many reports of healthy individuals successfully controlling SARS-CoV-2 infection without having detectable neutralising, or receptor-binding domain (RBD), antibodies while having prominent SARS-CoV-2-specific T-cell memory^2,7-10^. Clinical interventions using monoclonal antibodies further suggest that humoral immunity to SARS-CoV-2, although important, is not the hoped-for panacea for individuals that require intensive treatment^11,12^.

The association of SARS-CoV-2-specific CD4 and CD8 T-cells with milder disease suggests that both T-cell subsets play a role in protective immunity^2^. Indeed, the relative scarcity of naïve T-cells in individuals over 65 years old and the connection between ageing and impaired adaptive immune responses to SARS-CoV-2 has been suggested as a major cause of severe disease^2^. Analysis of immune cells in bronchoalveolar fluid from patients with COVID-19 showed that moderate disease correlated with highly clonally expanded CD8 T-cells^13^. Acute phase SARS-CoV-2-specific T-cells displaying a highly activated cytotoxic phenotype were present in antibody-seronegative exposed family members indicating that they may be capable of eliminating infection prior to induction of humoral immunity^7^ and it has been suggested that strong antibody responses but weak CD8 T-cell responses could contribute to acute COVID-19 pathogenesis and severity^1,14^. Depletion of CD8 T-cells in convalescent non-human primates reduced the protective efficacy of natural immunity against SARS-CoV-2 rechallenge^15^ suggesting CD8 T-cells in the upper respiratory tract may play a similar protective role in humans. Given the importance of CD8 T-cells to adaptive immune protection against COVID-19, we set out to examine dominant T-cell responses to SARS-CoV-2 through the most prevalent major histocompatibility complex (MHC) allele in humans, *HLA A*02* ^16^. HLA A*02 is an MHC class I molecule and presents processed intracellular protein antigens at the cell surface in the form of short peptides 8-10 amino acids in length for inspection by CD8 T-cells.

SARS CoV-2 infection induces T-cells that recognise peptides derived from a range of viral proteins with enrichment for those that respond to Spike, nucleocapsid, membrane, ORF1ab and ORF3a^7,8,17-19^. Unbiased screening of nine *HLA A*0201*^+^ convalescent patients (CP) showed that the two biggest and most frequent CD8 T-cell responses recognised regions of the virus contained within ORF1ab residues 3,881-3,900 and Spike residues 261-280^19^. The dominant Spike epitope was narrowed down to residues 269-277 (YLQPRTFLL) and T-cells that responded to this peptide represented >1 in 10,000 total CD8 T-cells in many *HLA A*0201*^+^ CP with the majority of the nine donors using a TCR made with the *TRAV-12-1* gene, suggesting a potential shared or ‘public’ response^19^. Another study found that YLQPRTFLL was the most frequently recognised of 13 reported HLA A*02:01-restricted epitopes (responses in 16/17 *HLA A*0201*^+^ CP studied)^10^. TCR sequencing of HLA A*02:01-YLQPRTFLL tetramer^+^ cells revealed prominent CDR3 motifs that were shared across individuals and confirmed the general *TRAV-12-1* dominance^9^. TCRβ repertoire analyses also indicates that CD8 T-cell responses to the Spike 265-277 region containing the YLQPRTFLL epitope dominate responses to Spike in both CP and individuals vaccinated with ‘single-shot’ Ad26.5.COV2.S vaccine in the US irrespective of HLA type^20^. We reasoned that if SARS-CoV-2 were to exhibit escape from CD8 T-cells then this would most likely first occur within a dominant T-cell response restricted by the most frequent HLA in the population. We therefore focussed our attention on residues 269-277 of the SARS-CoV-2 Spike protein and found that the most prevalent mutation in this T-cell epitope, YLQ**<u>L</u>**RTFLL (P272L change indicated in bold underlined text), was not recognised by any of the >120 TCRs that responded to the founder epitope (YLQPRTFLL) across a cohort of nine HLA A*02^+^ CP. We also found sizeable populations of CD8 T-cells that stained with peptide-HLA A*02 multimers bearing the YLQPRTFLL peptide in a cohort of individuals that had been vaccinated against SARS-CoV-2. These cells could not be stained by reagents manufactured with the P272L variant suggesting that this variant escapes from vaccine induced T-cell responses.

## RESULTS

### Emergence of mutations in the YLQPRTFLL CD8 T-cell epitope

The previously unprecedented genome-sequencing efforts for SARS-CoV-2 and sequencing of over 400,000 genomes in the United Kingdom to date (>45% of the global effort) has allowed detailed analyses and identification of over a thousand UK transmission lineages, with some variants associating with higher viral loads and increased transmission fitness^21,22^. We examined the global dataset sequenced as of January 31, 2021 and performed focussed analyses on the dominant Spike epitope at residues 269-277 (**Figures 1, 2** and **Supplementary Figures 1**). Within our dataset, we used ancestral state reconstruction to estimate the number of independent amino acid substitutions that have occurred in this region from sequenced data collected from the beginning of the pandemic to January 31, 2021. This analysis identified at least 24 different amino acid changes in this region, 22 of which were seen six or fewer times. The two variants resulting in the largest number of cases both occurred at position 272 (P272L and P272H). The P272H mutation occurred prior to onward transmission that resulted in 26 sequenced cases while P272L prior to onward transmission that led to an international cluster comprising over a thousand sequenced cases (**Figure 2** and **Supplementary Figure 1**). The P272L mutation occurred on a background of the B.1.177 lineage (**Figure 1**). B.1.177 is characterised by the lineage defining Spike mutation A222V and was shown to have arisen in Spain, and to have been exported to Europe and the wider world through travel over the summer of 2020^23^. B.1.177 was responsible for numerous local outbreaks in the autumn of 2020 in the UK and was a key lineage in establishing the ‘second wave’ of SARS-CoV-2 in the UK and Europe. The genomic data indicates that the first sequenced occurrence of P272L in any lineage was identified in March 2020, occurring in lineages B1 and B.1.1.263 before emerging in the B.1.177 background, with the first sequenced case of B.1.177 with P272L being reported in June 2020. Following its emergence, P272L in B.1.177 has been sequenced from cases in 29 countries, implying extensive spread over the summer and autumn of 2020 (**Figures 1&2** and **Supplementary Figure 1 and 2**). The B.1.177 sub-lineage carrying P272L was likely to have been imported into the UK in June, shortly after it was first observed (**Supplementary Figure 1**) and has spread extensively since then becoming the 4^th^ most frequent Spike variant in B.1.177 (**Figure 1**). Predominance of the B.1.177 P272L variant was higher in the South East of the UK and was present in 678/4,317 (∼16%) of all B.1.177 genomes sequenced from Greater London between March 7 2020 and January 31 2021. Although this lineage was ultimately suppressed by non-pharmaceutical interventions in the UK following the identification of B.1.1.7, up until December 2020 B.1.177 with P272L showed growth in multiple parts of the UK (**Figure 1**). Although global frequency counts are skewed by the emergence of variants such as B.1.1.7 and variable sequencing globally, in absolute terms P272L was in the top 15 Spike mutations observed globally as of January 31, 2021 (**Supplementary Figure 2A**) and by that point the variant had arisen independently at least 4 further times in other lineages (**Figure 1C**). As of July 2021, P272L has independently emerged, and been sequenced twice or more, in at least 13 lineages other than B.1.177 and its sublineages (**Supplementary Table 1**). It is also notable that this mutation has arisen spontaneously in variants of concern including B.1.1.7/Alpha B.1.351/Beta (9 sequenced cases of Beta with P272L in public databases as of the of July 13, 2021) and has also been seen in at least one case from each of the B.1.617.2/Delta and P.1/Gamma backgrounds, as well as being present in backgrounds that are potential variants of interest/concern including B.1.429 (13 sequenced cases) and B.1.526 (5 sequenced cases). In the case of B.1.1.7, P272L has been observed in 246 sequenced cases of this lineage as of May 31, 2021 (**Supplementary Figure 2E**). 32% (71/220) of reported sequences in the period Feb-June 2021 were in Campania, Italy (peaking in April 2021) with other local outbreaks of P272L-carrying B.1.1.7 evident in Nebraska/Georgia USA and the UK (∼29% and ∼14% of total P272L in this lineage Feb-June 2021). The geographic and temporal clustering of P272L within sequenced B.1.177 and B.1.1.7 lineages is suggestive of localised transmission.

**Figure 1.**
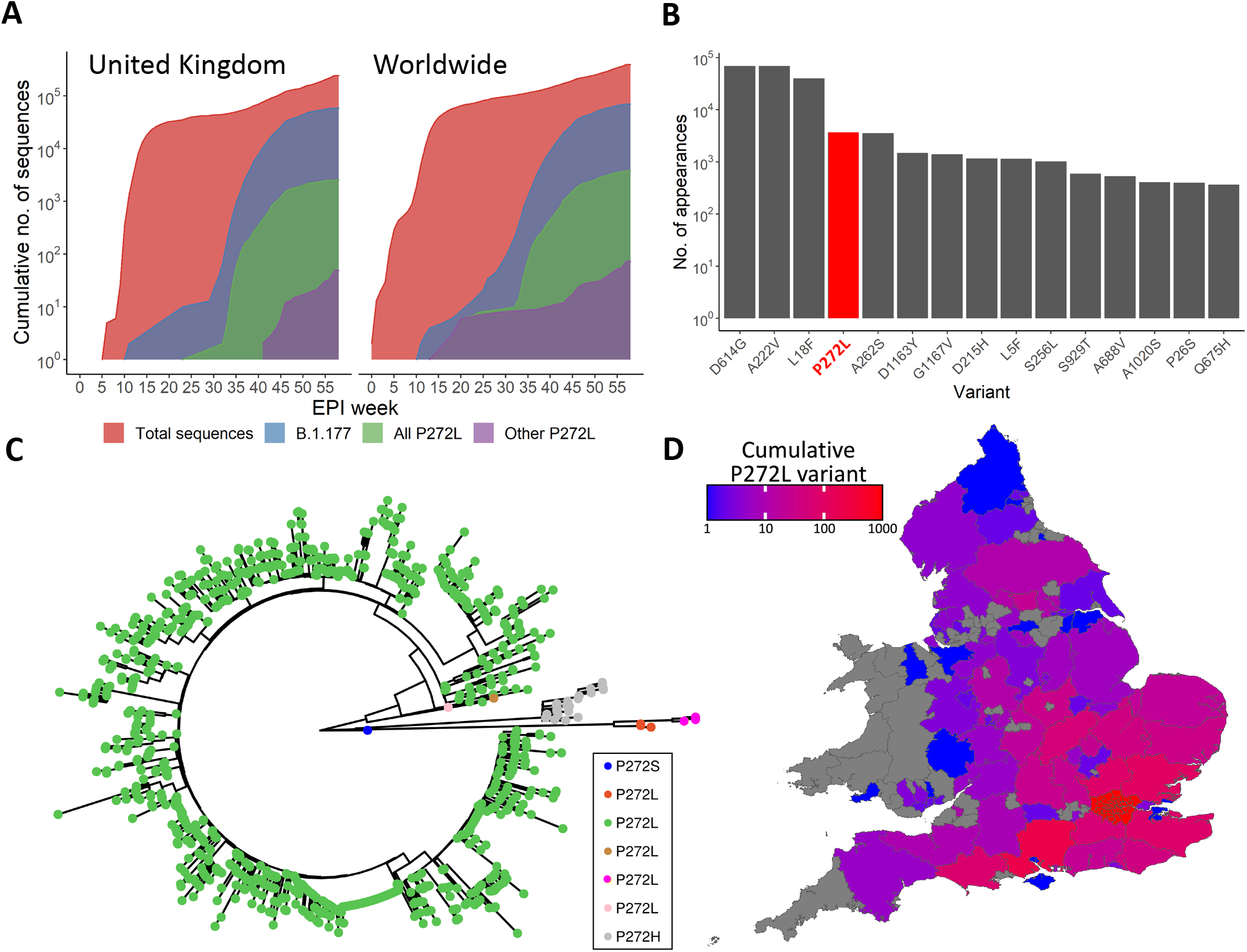
SARS-CoV-2 variation in the immunodominant Wuhan-YLQPRTFLL dominant HLA A*02:01-restricted CD8 T-cell epitope. (**A**) Cumulative frequency of all sequences; sequences in the B.1.177 lineage; and sequences possessing the P272L variant, both in B.1.177 and other lineages across the United Kingdom (left) and worldwide (right). Total number of P272L variants by nation, binned into periods of 10 epidemiological (EPI) weeks is shown in Supplementary Figure 1. **(B**) Top 15 most frequently observed variants observed in worldwide Spike glycoprotein sequence data in lineage B.1.177. Data for all lineages is shown in Supplementary Figure 3. **(C**) Phylogenetic tree showing 1227 taxa with variants at Spike position 272, with colours indicating subtrees representing potential independent mutations, computed by ASR on a larger tree of 200 221 taxa (not shown). **(D**) Total number of sequences possessing P272L variant in England and Wales per administrative region up to and including 31^st^ January 2021.

**Figure 2.**
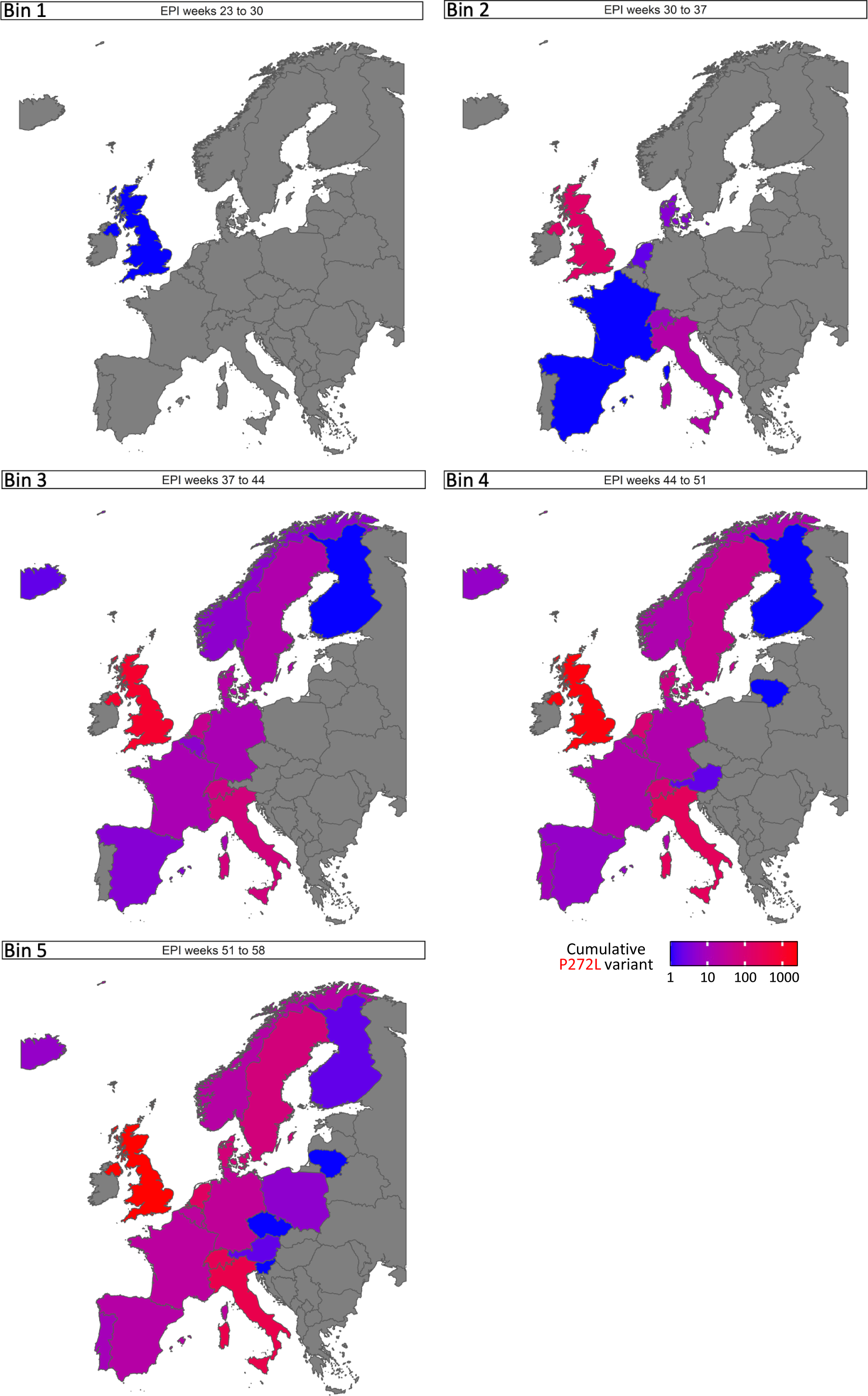
SARS-CoV-2 viral dynamics of P272L spike mutation in Western Europe. Total number of P272L variants by nation, binned into periods of 8 epidemiological (EPI) weeks, from EPI week 23 (beginning Sunday 31^st^ May 2020), the first time a sequence with P272L was observed in sequence data; up to and including January 31^st^ 2021 (EPI week 58).

### The P272L Spike variant is not recognised by YLQPRTFLL-specific CD8 T-cells from convalescent patients

In order to study the impact of mutation in the YLQPRTFLL epitope we recruited a cohort of SARS-CoV-2 convalescent local healthcare workers during June 2020. Each had a history of COVID-19 symptoms and a positive nasopharyngeal swab for SARS-CoV-2 by PCR >28 days prior to sample collection. 15/30 donors tested (50%) were *HLA A*02*^+^ by antibody staining. Nine of the first ten *HLA A*02*^+^ CP we screened had CD8 T-cells that stained with A*02:01-YLQPRTFLL tetramer (example in **Figure 3A**), confirming the widespread response to this epitope seen in previous studies^10,19^. Bulk sorting of tetramer^+^ populations was used to generate a T-cell line from each of the nine CP initially screened. These lines all responded to YLQPRTFLL peptide (13-53% total cells) but completely failed to respond to YLQ**<u>L</u>**RTFLL peptide (**Figure 3B and Supplementary Figure 3A)** despite this sequence showing improved binding to HLA A*02 compared to the parental (Wuhan) sequence (**Supplementary Figure 3B**). The ability of the YLQ**<u>L</u>**RTFLL peptide to bind to HLA A*02 was confirmed during tetramer production (**Supplementary Figure 3C**). TCR sequencing of the A*02:01-YLQPRTFLL tetramer^+^ T-cells in each line identified 121 different TCRs across the seven patients including the public TCR chains previously identified in addition to many donor-specific TCR sequences (**Figure 3C** and **Supplementary Figure 4**). Remarkably, these data indicate that all the TCRs failed to respond to the P272L variant sequence and suggests that the proline at residue 272 plays a critical role in recognition by all TCRs across this cohort. Failure to recognise the P272L mutant or bind to this sequence was confirmed by direct *ex vivo* staining of PBMC (**Figure 3A**) and by using four different CP-derived T-cell clones (**Figure 4**). We further confirmed that surrogate infected cells expressing full-length Spike protein with the P272L substitution (**Supplementary Figure 5**) were not recognised by any of these T-cell clones (**Figure 4**). We conclude that P272L has arisen on multiple occasions (appearing to show selection in the B.1.177 background) and escapes from all TCRs raised against the parental (Wuhan) sequence that is being used in all current vaccines.

**Figure 3.**
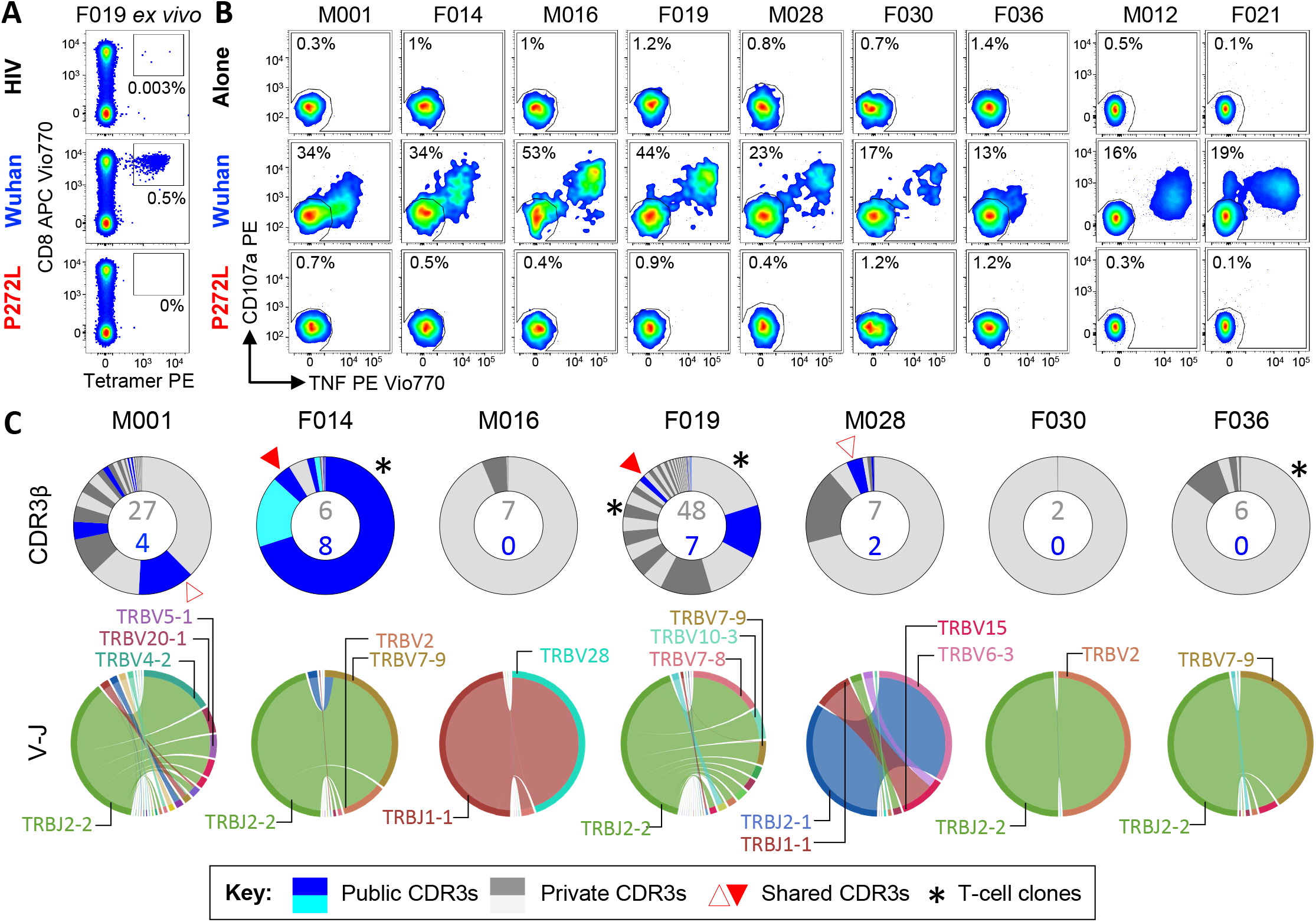
Wuhan-YLQPRTFLL T-cell lines from SARS-CoV-2 positive donors do not activate with P272L-YLQ<u>L</u>RTFLL variant peptide. **(A**) *Ex vivo* PBMC Wuhan-YLQPRTFLL and P272L-YLQ**<u>L</u>**RTFLL tetramer staining of a COVID patient. HLA A2 SLYNTVATL (SLY) peptide from HIV used as an irrelevant tetramer^44^. Percentage of CD8 T-cells is shown. **(B**) Wuhan-YLQPRTFLL T-cell line enriched from nine COVID patients do not activate towards the P272L-YLQ**<u>L</u>**RTFLL peptide (10^−6^ M used for both peptides). Percentage of reactive cells is displayed. Full data shown graphically in **Supplementary Figure 3A**. M012 and F021 analysis performed on a different day. **(C**) T-cell receptor (TCR) beta chain analysis of Wuhan-YLQPRTFLL tetramer sorted T-cells from seven of the patients in A (flow data in **Supplementary Figure 4A**). Pie charts display the proportion (chart segments) and frequency (numbers in center) of public (blue) and private (grey) TCRs. Variable (V) (arc on the right) and joining (J) (arc on the left) gene rearrangements are shown below the pie charts, with the dominant clonotypes annotated. CDR3s sequences are listed in **Supplementary Figure 4B**. T-cell clones (indicated with asterisk) are featured in **Figure 4**.

**Figure 4:**
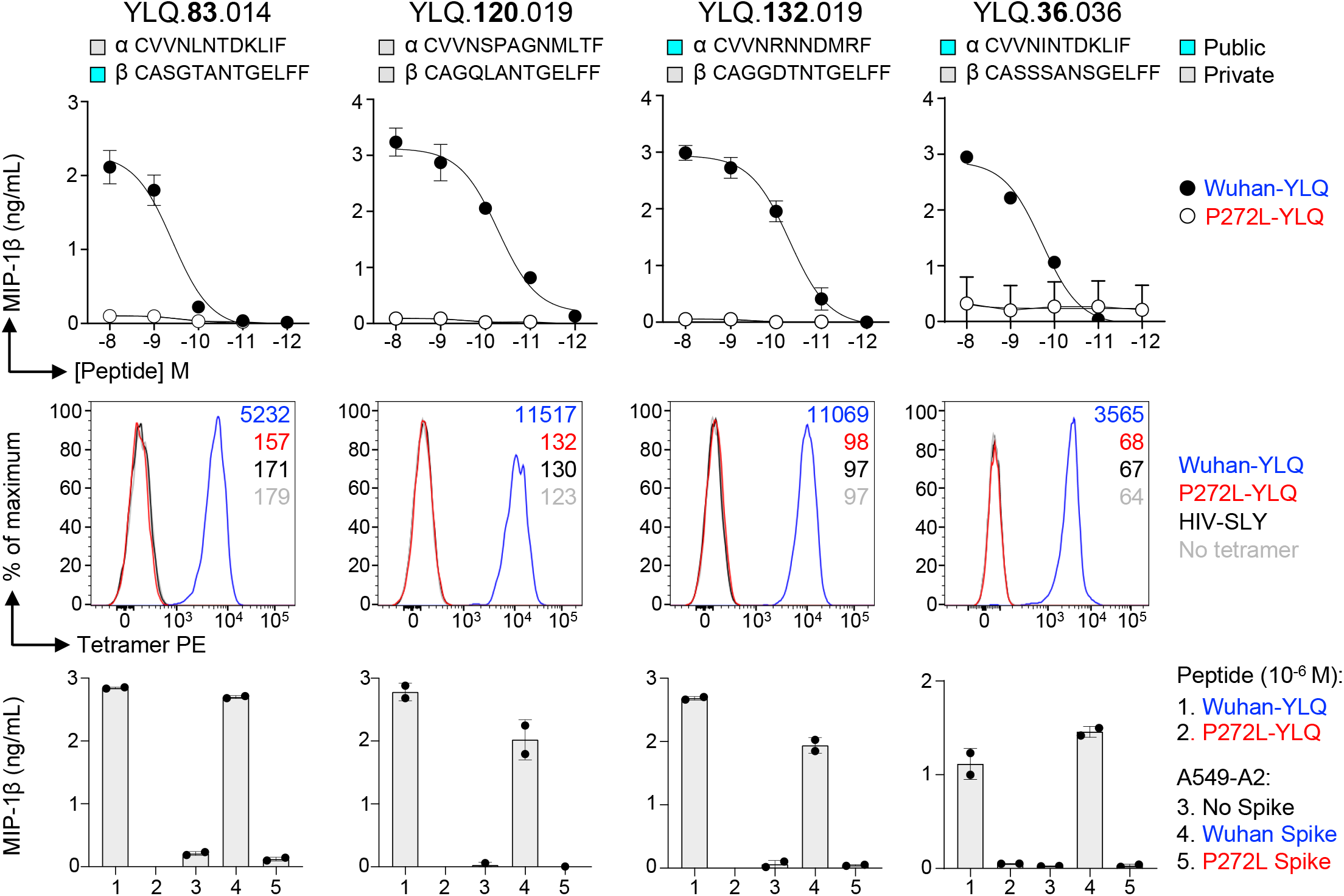
Wuhan-YLQPRTFLL specific T-cell clones from SARS-CoV-2 convalescent patients are unable to recognize P272L variant. YLQPRTFLL peptide reactive CD8 clones were grown from donors F014 (83), F019 (120 and 132) and F036 (36). Alpha and beta TCR chain CDR3s of each clone are shown and the public or private status of each indicated according to the key. Upper panel: peptide sensitivity assay with Wuhan-YLQPRTFLL and P272L-YLQ**<u>L</u>**RTFLL peptides. MIP-1β ELISA and error bars depict SD of duplicates. Middle panel: peptide-HLA tetramer staining. HLA A2 SLYNTVATL (SLY) peptide from HIV used as an irrelevant tetramer^41^. Lower panel: A549 cells expressing HLA A2 (A549-A2) and full length P272L Spike were not recognized, whereas endogenously expressed Wuhan spike was recognized. MIP-1β ELISA and error bars depict SD of duplicates.

### The P272L Spike variant is not recognised by YLQPRTFLL-specific CD8 T-cells from vaccinees

We next examined responses to YLQPRTFLL in individuals who had been vaccinated against SARS-CoV-2. In total, PBMC from four HLA A*02^+^ healthy donors (HD) who had received a single dose of the AstraZeneca ChAdOx1 nCoV-19 vaccine^24^ and a further three HD who received the Pfizer-BioNTech BNT162b2 vaccine^25^ (see Materials and Methods for donor and vaccination details) were stained with peptide-HLA tetramers. All vaccinees had HLA A*02-YLQPRTFLL tetramer staining T-cells that accounted for between 0.01 and 0.2% of total CD8 T-cells (**Figure 5**). In all cases, these T-cells failed to stain with the HLA A*02-YLQ**<u>L</u>**RTFLL (P272L) reagent in parallel assays, indicating that the TCR on these T-cells did not strongly engage this sequence in accordance with the CP cohort data (**Figure 5**). We conclude that the YLQPRTFLL-specific T-cells induced by two current COVID vaccines fail to engage to the P272L variant sequence.

**Figure 5:**
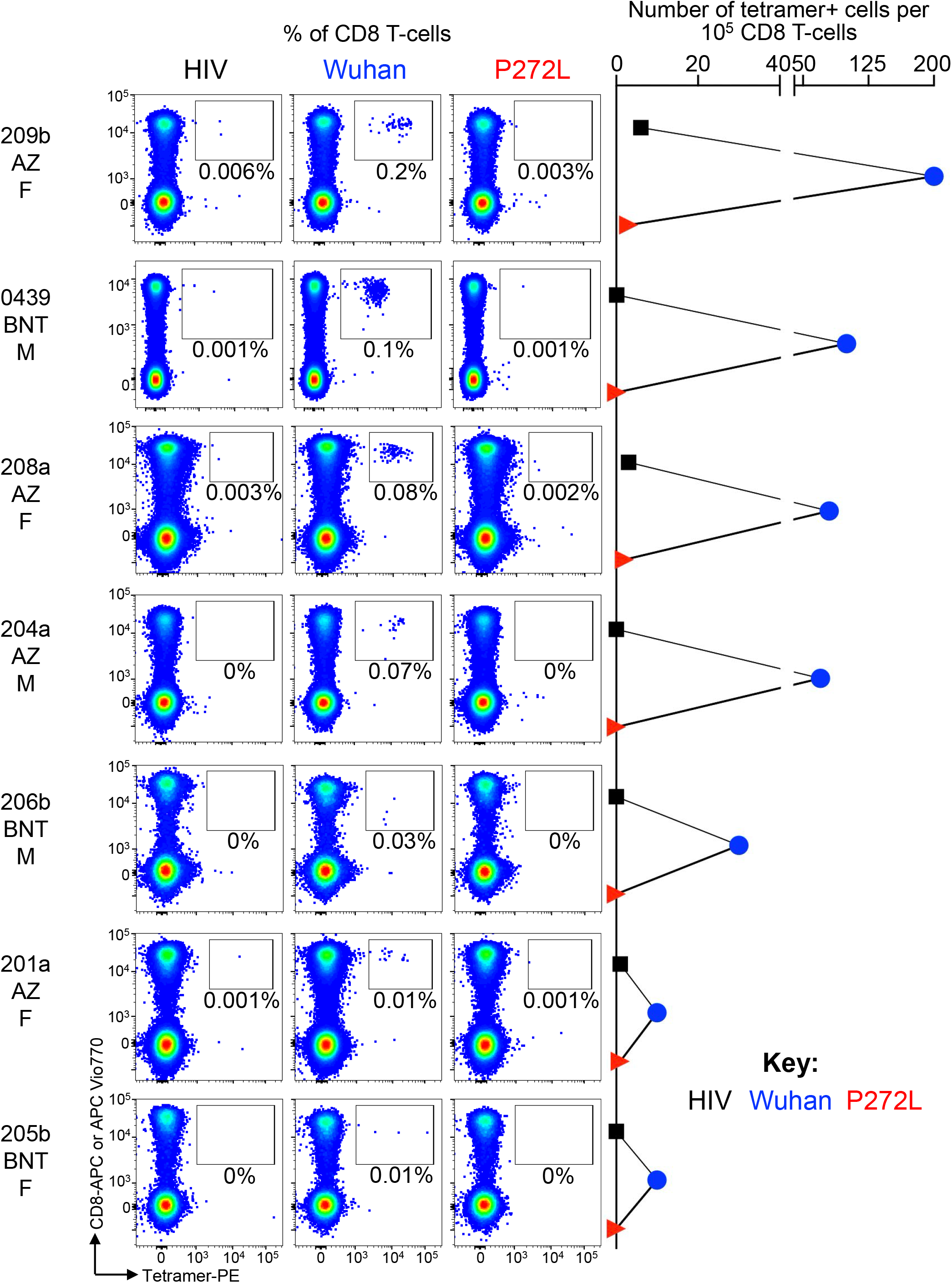
Wuhan-YLQPRTFLL specific T-cell clonotypes from vaccinated donors fail to stain with P272L variant tetramers. Tetramer staining of PBMC from 7 vaccinated donors (six single dose and donor 0439 double dose) with HLA A*0201-YLQPRTFLL and P272L tetramers. Donors received the AstraZenica (AZ) ChAdOx1 nCov-19 vaccine or the BNT162b2 (BNT) vaccine as indicated. See materials and Methods for details of each donor. The age and gender indicated under donor name. 0439 BNT: BD FACS Canto with CD8-APC-Vio770. All other donors: SONY MA900 with CD8 APC.

### Structural comparison of HLA A*02:01-YLQPRTFLL and HLA A*02:01-YLQ<u>L</u>RTFLL

T-cells are known to be widely crossreactive, allowing some TCRs within polyclonal antigen-specific T-cell populations to recognise viral mutations presented at the surface of infected cells in the context of HLA-I molecules^26^. Indeed, individual T-cell clonotypes that exhibit greater crossreactivity with known, naturally arising viral variants at an immunodominant CD8 T-cell epitope have been suggested to protect against disease progression in HIV infection^27^. We found it surprising that all 121 HLA A*02-YLQPRTFLL-specific TCRs in our CP cohort failed to recognise the P272L variant and that this variant failed to stain T-cells in all vaccinees we studied. In order to understand why this single amino acid substitution had such a drastic effect on recognition by many TCRs we solved and compared the structures of YLQPRTFLL and YLQ**<u>L</u>**RTFLL bound to HLA A*0201 at 1.67 Å and 2.0 Å respectively (**Figure 6)**. The HLA A*0201-YLQPRTFLL structure showed the Arginine at position 273 forms an obvious protruding positively charged focal point for TCRs that is stabilized by six intrapeptide bonds including three hydrogen bonds (**Figure 6A**). The HLA A*0201-YLQ**<u>L</u>**RTFLL structure shows fewer internal contacts that maintain the shape and dimensions of the bulged part of the epitope (**Figure 6B**). The 2.7 to 3.5 Å further extension of the amino acid 272 side chain in the P272L variant compared with the parent (Wuhan) epitope is likely to sterically interfere with TCRs that make contacts with the substituted proline, although TCR-peptide-HLA structures will be required to formally confirm this supposition (**Figure 6C**).

**Figure 6:**
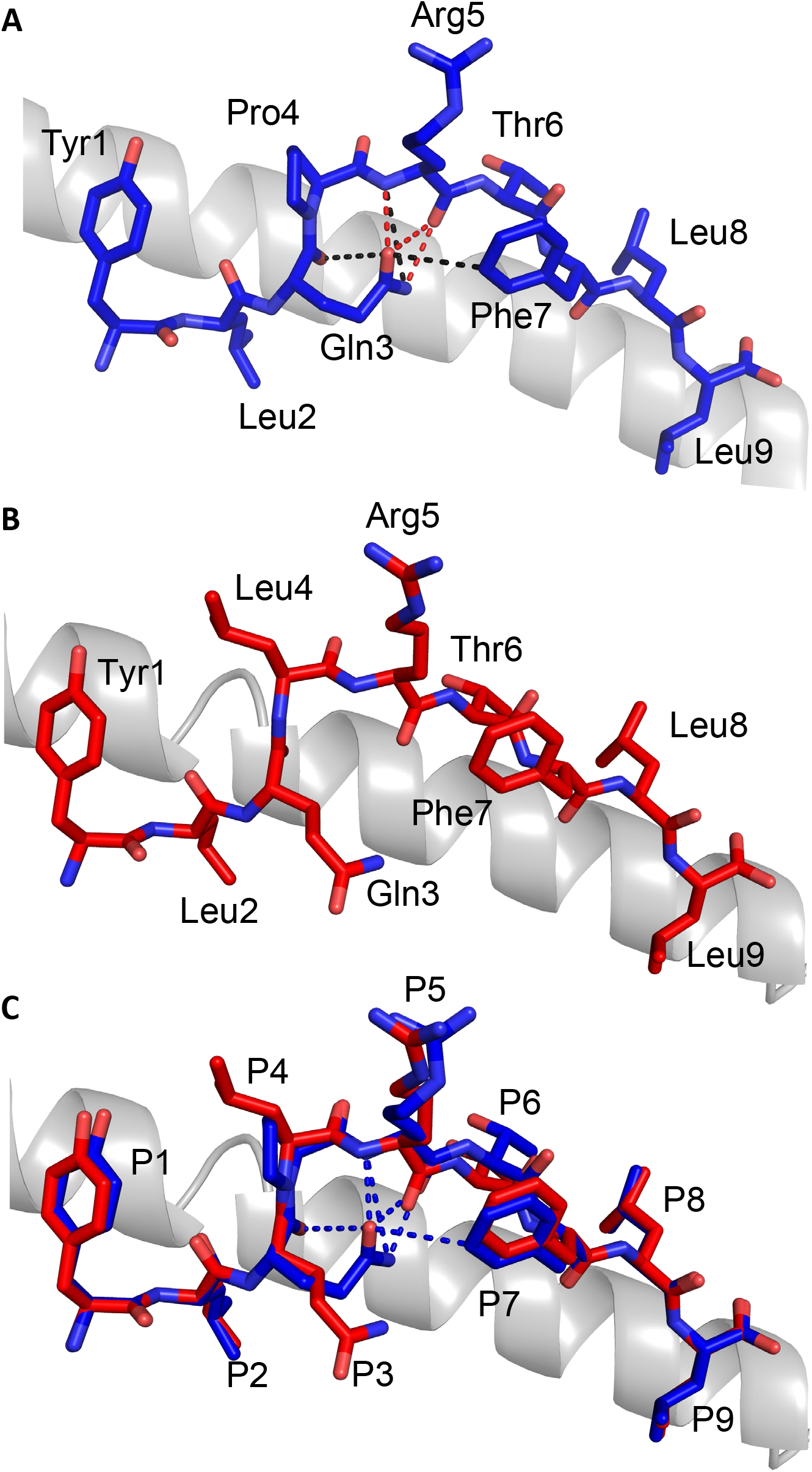
Structural differences in HLA A*02 presenting Wuhan-YLQPRTFLL and P272L epitopes. (**A**&**B**) 3D structures of HLA A*02-YLQPRTFLL (**A**) and HLA A*02-YLQ**<u>L</u>**RTFLL (**B**). Peptides shown as sticks. HLA A*02 shown as grey cartoon. Van Der Waals interactions and hydrogen bonds shown as black and red dashes respectively. (**C**) A comparison between HLA A*02-YLQPRTFLL (blue) and HLA A*02-YLQ**<u>L</u>**RTFLL (red) peptide presentation. Peptides shown as sticks. HLA A*02 shown as grey cartoon. Interactions within the YLQPRTFLL peptide are shown as blue dashes. Data collection and refinement statistics are shown in Supplementary Table 1 and electron density maps around the peptides in the MHC structures are shown in Supplementary Figure 6.

## DISCUSSION

Emerging evidence indicates that HLA-I-restricted CD8 ‘killer’ T-cells contribute to the control of SARS-CoV-2 and the immunity to disease offered by the currently approved vaccines^1,2^. Individual HLA-I alleles can be associated with increased likelihood of control or progression of disease with viruses such as HIV^28^. The immune pressure of prevalent host CD8 T-cell responses against HIV results in selection of altered viral sequences and viral immune escape^29-31^. We were interested in whether there was evidence that SARS-CoV-2 might also escape from CD8 T-cells. Genome sequencing has shown extensive non-synonymous mutations occurred within the SARS-CoV-2 genome during 2020. Some of these mutations alter viral fitness^32^ while others have been shown to diminish the binding of antibodies^33^. To date, there has been limited study of T-cell escape and a broad brush-approach of examining T-cell recognition of key transmission variants failed to reveal any evidence that escape was occurring^34^. A recent report demonstrated that mutations in the RBD that enhance angiotensin converting enzyme (ACE)2 binding and viral infectivity concomitantly reduce T-cell recognition through HLA*24:02 ^35^. Another study showed that some SARS-CoV-2 mutations in HLA-I-restricted epitopes resulted in lower binding to the HLA leading to reduced recognition by CD8 T-cells but there was little evidence of these mutations being disseminated^36^. It has been suggested that the L270F (Y**<u>F</u>**QPRTFLL) variant might be an escape mutant^36^, but this variant was only seen six times throughout our study period, in six different locations and five different PANGO lineages. From our data, this would suggest that while this mutation does arise, there is no evidence of spread of this variant following sporadic emergence across multiple different SARS-CoV-2 lineages.

We reasoned that if escape were to occur then it would be most likely to be seen in a prevalent response through a frequent HLA. HLA A*02 is frequently carried by all human populations except those of recent African ancestry where it is present at lower levels. HLA A*02 is believed to be the most frequent HLA-I in the human population worldwide occurring in ∼40% of individuals^16^. *HLA A*02* is hypothesized to have become so frequent in the population due to its success at presenting well-recognised epitopes from historically dangerous pathogens, a supposition consistent with this HLA having entered the population via interbreeding with Neanderthals and Denisovans after early humans migrated from Africa^37^. A previous study used overlapping peptides from the entire SARS-CoV-2 proteome to identify that the most common HLA A*02-restricted responses in CP were to epitopes contained within peptides spanning residues 3,881-3900 of ORF1ab and 261-280 of Spike^19^. The Spike epitope was narrowed down to residues 269-277; sequence YLQPRTFLL. A further study confirmed the prevalence of CD8 T-cells that respond to YLQPRTFLL in CP that are absent in healthy donor blood taken before the spread of SARS-CoV-2 ^10^. We confirmed the prevalence of responses to this epitope in our cohort where responses were observed in 9/10 HLA A*02^+^ CP tested and all 7/7 SARS-CoV-2 vaccinees including those who had received only the first dose of a double dose schedule.

We hypothesised that if escape from CD8 T-cells were to occur, then it would most likely be seen in a dominant epitope restricted by the most frequent HLA-I in the population so focussed our attention on nonsynonymous mutations in the sequence encoding YLQPRTFLL. While at least 24 different amino acid changes have occurred in this region, only two were seen in more than six sequenced cases in the period from the beginning of the pandemic to January 31^st^, 2021. The two variants resulting in the largest number of cases both occurred at position 272 with the P272L mutation occurring in over 8,500 sequenced cases as of June 1^st^, 2021. The P272L variant arose in 5 different lineages during the study period but was most prevalent in the B.1.177 lineage that played a key role in establishing the ‘second wave’ of SARS-CoV-2 in the UK and Europe during the Autumn of 2020. P272L was the 4^th^ most common variant seen in the B.1.177 lineage and its descendants and in the top 15 Spike mutations observed globally to January 31, 2021. Nine of ten of our HLA A*02^+^ CP cohort tested made a response to the YLQPRTFLL epitope; a response comprising >120 different TCRs in total. Remarkably, no response was seen to the P272L variant. This finding was confirmed using four CP-derived T-cell clones that included TCR α or β chains that have previously been described in other CP cohorts^10,19^. Responses to the YLQPRTFLL epitope in a cohort of HLA A*02^+^ healthy donors who had been vaccinated against SARS-CoV-2 but had never had symptoms or a positive test since January 2020 ranged from 0.01-0.2% of CD8 T-cells by peptide-HLA staining. However, all of the T-cells raised against the YLQPRTFLL epitope across our vaccinee cohort expressed TCRs that engaged the P272L very poorly and did not stain with P272L peptide-HLA tetramers. The fact that >120 different TCRs raised against the YLQPRTFLL epitope all fail to see the P272L variant suggests that either the proline at position 272 must form a major focus of TCR contact that a leucine residue cannot compensate for or that substitution with leucine at this position must interfere with a commonly shared TCR binding mode (or both). Comparison of the atomic structures of HLA A*02-YLQPRTFLL and HLA A*02-YLQ**<u>L</u>**RTFLL showed a largely preserved fold, but with local geometry alterations at the epitope residues 3 to 5 (Spike residues 271-273) which presumably form the majority of contacts with TCRs. However, a full understanding of how these changes impact on TCR binding will require TCR-peptide-HLA structures.

SARS-CoV-2 variants have been rapidly emerging throughout the current pandemic with mutants that enhance transmissibility, evade host immunity or increase disease severity being of particular concern. Current systems for identifying variants of public health concern involve risk assessments that identify mutations that are present, and then looks for mutations that have a known biological effect in order to assess their significance. While extensive amounts of work have been undertaken to predict and characterise the likely effects of mutations in Spike on antibody recognition, the same is not true for T-cells. Current risk assessments that only consider antibody affinity and factors associated with transmissibility (e.g. mutations that improve cellular infectivity) are clearly incomplete. The range of potential effects of mutations that affect recognition by T-cells is significant, and urgently requires further study. It is clear that there was a significant outbreak that spanned Europe of a lineage carrying a mutation that we have shown has a detectable impact on T-cell recognition, which went unrecognised at the time. Ultimately, it is unclear whether SARS-CoV-2 incorporates enough genetic plasticity to allow it to escape from humoral immune responses. However, the area targeted on the RBD by neutralising antibodies is large enough, and the range of epitopes targeted in polyclonal human sera broad enough, to ensure that no single mutation should allow complete escape from neutralisation in the majority of individuals^38^. Likewise, the wide array of HLA across the population^16^ and the broad range of epitopes responded to in COVID-19 patients^18^ combine to make it unlikely that SARS-CoV-2 will completely escape from T-cell surveillance in the near future. During the early stages of the pandemic, it seems likely that alterations in SARS-CoV-2 Spike that enhance binding to ACE2 or reduce the capability of vaccine-induced, or natural, antibodies to neutralise the virus will take precedence due to their potential relevance in all transmission events. This ‘shrouding effect’ may well have occurred in the UK SARS-CoV-2 epidemic, although this is difficult to fully assess for two reasons. First, the B.1.1.7/Alpha lineage has been shown to have a significantly higher transmissibility compared to B.1.177 (and other circulating lineages occurring in the Autumn and Winter of 2020)^39^, and so while P272L may have conferred some advantage, this is likely to have been insufficient to prevent the lineage being replaced by B.1.1.7/Alpha. Also, it is important to note that the trajectory of P272L B.1.177 isolates was generally upwards going into the Winter of 2020, however the introduction of significant non-pharmaceutical interventions in response to B.1.1.7/Alpha resulted in a strong suppression of all SARS-CoV-2 lineages in the UK. It remains to be seen whether the P272L mutant will arise and increase in relative frequency in other lineages. However, this mutation has occurred in several variants already, with sequenced cases carrying this mutation from three variants of concern (Alpha, Beta and Delta) all existing in public sequence databases as of July 2021. Amongst these, numbers were highest in Alpha/B.1.1.7, with the sequence data suggesting some localised spread of P272L B.1.1.7/Alpha in early 2021 prior to emergence and replacement of B.1.1.7/Alpha by B.1.617.2/Delta in Europe and elsewhere. Based upon this observation, we predict that it will appear in future variants, particularly as global vaccination rates increase. Our data suggest that the 269-277 epitope of Spike is one region that should be monitored and should feature in risk assessments by public health agencies going forward.

In summary, we demonstrate that SARS-CoV-2 can readily alter its Spike protein via a single amino acid substitution so that it is not recognised by CD8 T-cells targeting the most prevalent epitope in Spike restricted by the most common HLA-I across the population. While it is not possible to directly attribute the emergence and propagation of the Spike P272L SARS-CoV-2 variant in parts of the UK where *HLA A*02* is frequently expressed to CD8 T-cell-mediated selection pressure, specific focussing of immune protection on a single protein (e.g. SARS-CoV-2 Spike favoured by all currently approved vaccines^40^) is likely to enhance any tendency for escape at predominant T-cell epitopes like YLQPRTFLL. Our demonstration that mutations that evade immunodominant T-cell responses through population-frequent HLA can readily arise and disseminate, strongly suggests that it will be prudent to monitor such occurrences and to increase the breadth of next generation SARS-CoV-2 vaccines to incorporate other viral proteins.

## METHODS

### SARS-CoV-2 lineage dynamics

To characterise the distribution of mutations in the Spike epitope between positions 269-277, we performed a set of analyses making use of high quality publicly available sequence data made available by GISAID and COG-UK as of 31 January 2021^42^. To generate an estimate of the number of times mutations have occurred within the epitope, we performed ACCTRAN^43^ ancestral state reconstruction using gotree (available at https://github.com/evolbioinfo/gotree), on a tree with 200 221 taxa, generated by COG-UK (COVID-19 Genomics UK (COG-UK), 2020), for each Spike protein site. Subtree groups were identified by nodes with a consensus to variant transition. The phylogenetic tree for visualising variants at position 272 comprises 1227 taxa, and was generated from an alignment of consensus genome sequences using mafft version 7.475, with ambiguous/problematic positions masked following^44^ using v4 of the problematic sites vcf (available here: https://github.com/W-L/ProblematicSites_SARS-CoV2/blob/master/archived_vcf/problematic_sites_sarsCov2.2020-12-22-11:15.vcf). The tree was built using IQ-TREE^45^, with a HKY+G model^46^, 1000 ultrafast bootstrap replicates^47^, and rooted using Wuhan isolate MN908947.3 as an outgroup. Tree drawing was performed with ggtree^48^. Information on case location (country, from GISAID and local authority from the COG-UK public data) and variant frequency by PANGO lineage was collated from GISAID and COG-UK and these were then visualised using R version 4.0.5.

### Convalescent COVID patients

All participant samples were pseudo-anonymised and the clinical Chief Investigator, Dr Lucy C Jones, was the only individual with access to the confidential record linking sample numbers to identifying information. Pseudo-anonymised sample numbers were not known to participants or to anyone outside of the research group. Our CP study was given a favourable opinion by the Health Research Authority (HRA) Research Ethics Committee London – (Brighton & Sussex), IRAS (Integrated Research Application System for HRA) 269506 and was also reviewed by Healthcare Research Wales and Cwm Taf Morgannwg University Health board, with research permissions granted.

Eligible participants were recruited during March-June 2020 and were adult (over 18) convalescent healthcare workers at Cwm Taf Morgannwg University Health Board. Inclusion criteria included a positive nasopharyngeal swab for SARS-CoV-2 by PCR, more than 28 days prior to blood sample collection. In addition to having had a positive SARS-CoV-2 swab test, all participants fulfilled the criteria for a definite diagnosis of COVID-19 at least 28 days prior to recruitment, defined by at least one of the following: new onset of pyrexia, a continuous cough, anosmia, or ageusia. ‘M’ and ‘F’ of the patient code denotes male or female, with 3 males and 7 females used for this study. Venous blood samples (50mls) were collected from 37 informed and consented participants. This was stored in EDTA for up to 3 hours and transported in UN3373 containers to the laboratory. Peripheral Blood Mononuclear Cells (PBMCs) were collected and frozen on the same day. All human blood was procured and handled in accordance with the guidelines of Cardiff University to conform to the United Kingdom Human Tissue Act 2004. HLA A*02 typing was performed by flow cytometry using anti-HLA A2 antibody (BB7.2, BioLegend, San Diego, CA, US). Of the nine *HLA A*0201*+ patients seven had mild disease and two had moderate disease in accordance with the US National Institutes of Health classification. None of the participants required treatment with invasive ventilation support. The 3 male and 6 female participants studied had ages ranging from 30 to 60 years and a median age of 50 years. Peripheral blood was collected between 6 and 13 weeks after the positive PCR swab (median 12 weeks).

### SARS-CoV-2 vaccinees

We recruited six participants who had received one dose of a SARS-CoV-2 vaccine as part of the National UK vaccination programme, under ethical approval (Central University Research Ethics Committee (CUREC), University of Oxford, Approval Reference: R71346/RE001). Inclusion criteria for participants included: no clinical history of symptoms of COVID-19 since January 2020, no history of a positive COVID swab or positive COVID antibody test. Exclusion criteria of participants included: immunological conditions which directly affect immune responses, current or recent medications which directly affect immune responses, recent history of a transfusion of blood products and pregnancy. Peripheral blood (50 mls) was collected between 28 to 60 days after receiving one dose of a vaccine for SARS-CoV-2 as indicated in **Table 1** and processed withing 4 hours of sampling. Participants received either the Astrazeneca (ChAdOx1 nCoV-19) vaccine or the Pfizer (BNT162b2 2) vaccine. Healthy donor 0439 was a colleague who received the BNT162b2 vaccine^25^ and volunteered a sample on day 14 following the second vaccine dose having heard that we were studying immune escape. Donor 0439 was the only donor to have undergone a full vaccination course as indicated in **Table 1**. The three male and four female donors had ages ranging from 41 to 75 years and a median age of 57 years.

**Table 1.**
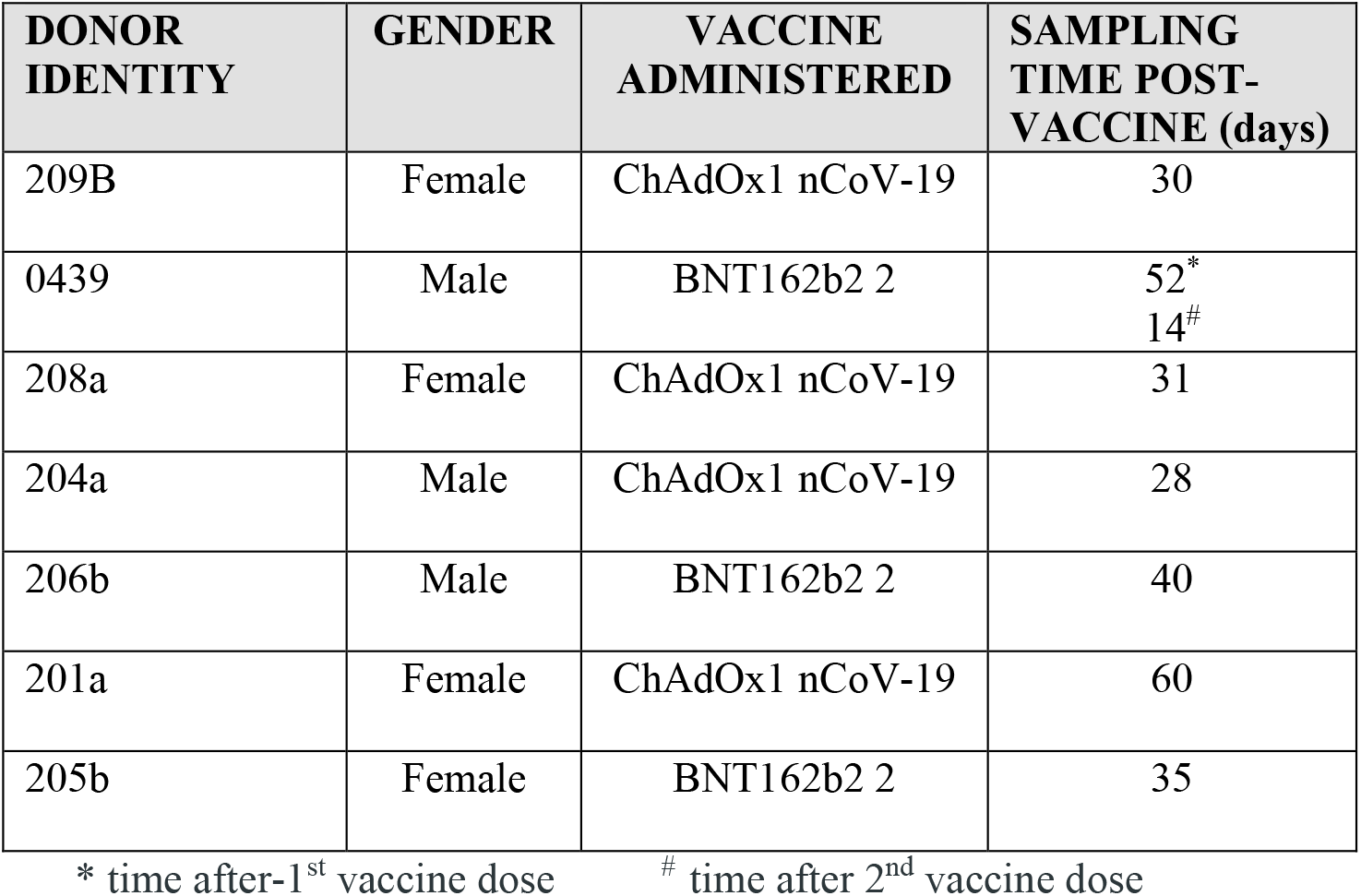
Participant characteristics. Gender, type of SARS-CoV-2 vaccine administered and the time of phlebotomy post-vaccine in days.

### PBMCs and cell lines

PBMC isolation from CPs and a healthy vaccinated donor was performed by standard density centrifugation using Histopaque-1077 (Merck Group, Darmstadt, Germany). Cell lines were cultured on the basis of ATCC guidelines: immortalized embryonic kidney cell HEK293T (CRL-1573™); lung carcinoma A549 (CCL-185 ™) and T2 (CRL-1992™). Cells were cultured in RPMI 1640 (A549 and T2) or DMEM (HEK293T) medium supplemented with 10% FBS, 2 mM L-glutamine and 100 U/mL penicillin and 100 μg/mL streptomycin (R10 or D10 respectively; all from Merck Group). Adherent cell lines were cultured as adherent monolayers, passaged when 50–80% confluent using D-PBS (Merck Group) with 2 mM EDTA to detach cells. T2s were cultured as suspension cells and split 1:10 twice weekly. Cells were regularly tested for mycoplasma contamination (MycoAlert™, Lonza).

### Generation of T-cell lines and clones

T-cells were cultured in R10 supplemented with 25 ng/ml interleukin IL-15 (Miltenyi Biotech, Bergisch Gladbach, Germany), 20 or 200 IU/mL IL-2 (Proleukin®; Prometheus, San Diego, CA), 1X non-essential amino acids solution, 1 mM sodium pyruvate and 10 mM HEPES buffer (all from Merck Group unless stated otherwise). T-cells from patients recognising Wuhan-YLQPRTFLL were isolated from whole PBMCs using PE conjugated Wuhan-YLQPRTFLL peptide-HLA tetramers and magnetic anti-PE beads according to the manufacturer’s guidelines (Miltenyi Biotec) and as previously described^49,50^. For donors M001, F012 and F021 a bulk T-cell library was firstly created using magnetically purified CD8 T-cells (Miltenyi Biotec) and CD3/CD28 Dynabeads according to the manufacturer’s instructions (ThermoFisher Scientific, Waltham, MA, US). After 2 weeks of expansion with CD3/CD28 beads, Wuhan-YLQPRTFLL PE tetramers and anti-PE beads were used as above to enrich antigen specific T-cells. Tetramer enriched cells were allowed to culture overnight and then expanded with irradiated (3100 cGy) allogeneic PBMCs from three donors purchased from the Welsh Blood Service, and 1 μg/mL of L-phytohemagglutinin (Pan Biotech, GmbH, Am Gewerbepark 6, 94501 Aidenbach, Germany). Cloning was performed as previously described^51^. Briefly, tetramer enriched T-cell lines were cloned by plating 0.5, 1 or 3 cells per well in 96U-well plates with 5 ×10^4^ irradiated PBMCs and PHA as above.

### Flow cytometry and cell sorting

For surface staining, typically 5 ×10^4^ cells per condition were used. T-cells were stained with LIVE/DEAD™ fixable violet dead stain (ThermoFisher Scientific) and CD3 (BW264/56) and CD8α (BW135/80) antibodies (all from Miltenyi Biotec). Cells were gated on FSC-A versus SSC-A, single cells (FSC-A versus FSC-H), then viable cells (marker of choice versus VIVID). Samples were acquired on a FACSCantoII machine (BD Biosciences, Franklin Lakes, NJ, USA) and analyzed in the FlowJo software (Tree Star Inc., Ashland, Oregon, US). Cell sorting was performed on SONY MA900 multi-application cell sorter (SONY, San Jose, CA, USA).

### Lentivirus production and transduction

Lentiviral particles were generated by transient co-transfection of HEK293T cells with a 3^rd^ generation plasmid system. Briefly, codon-optimised Wuhan Spike, P272L Spike (site directed mutagenesis of Wuhan Spike, details below) and *HLA A*0201* genes were synthesised (Genewiz, Essex, UK) and cloned into a 3^rd^ generation lentiviral plasmids. Spike genes were designed with a rCD2 co-marker: Xba1-Kozak-Spike-Xho1-P2A-rCD2-Sal1-Stop, and cloned in to pSF (Oxgene, Oxford Genetics Ltd, Littlemore, Oxford, UK). The pSF plasmid was modified by removal of a Xho1 site present in the plasmid backbone and insertion of stop codons after the Sal1 site. *HLA A2* was cloned in to pELNS (Kind gift from James Riley): Xba1-Kozak-HLA A2-Stop-Xho1. For transfection, pSF or pELNS (1.52 μg), envelope plasmid (pMD2.G; 0.72 μg) and packaging plasmids (pMDLg/pRRE; 1.83μg and pRSV-REV; 1.83μg) were mixed in 300 μL of Opti-MEM, reduced serum medium (ThermoFisher Scientific) followed by mixing with 1 μg/μl Polyethylenimine (PEI; Merck Group) at a 3:1 PEI: plasmid ratio. Plasmid/PEI mixtures were incubated at room temperature for 15 min, added dropwise to the HEK293T cells (80% confluence in one well of a 6-well plate) and incubated at 37°C in a 5% CO_2_ humidified atmosphere. The supernatants containing lentiviral vectors were harvested, 0.4 μm filtered, and used immediately for transduction or stored at -80°C and only defrosted once before transduction. A549 cells were transduced by spinfection with viral supernatant at 400g for 2 h, incubated at 37°C overnight and media was replaced the following morning. HLA A2 transduction was confirmed by staining with BB7.2 Ab. Spike transduced cells were firstly checked for with conjugated anti-rCD2 antibody (OX-34, BioLegend), then in follow-up experiments with 1 μg/test of unconjugated anti-SARS Spike glycoprotein antibody (1A9, Abcam, Cambridge, MA, US). An unconjugated isotype matched antibody (P.3.6.2.8.1, eBiosciences, ThermoFisher Scientific) was used as a control. A goat anti-mouse secondary antibody (multiple adsorbed PE conjugated Ig polyclonal; BD Biosciences, Oxford, UK) was used to detect Spike antibody staining.

### Site directed mutagenesis of the Wuhan Spike gene

Oligonucleotides were designed to substitute the Proline to a Leucine at position 272. Non-overlapping sequences were designed so that the 5’ end of the forward oligo (5’ TGAGAACCTTCCTGCTGAAGTACAACG 3’) started with the nucleotides to be mutated and included at least 20 complementary nucleotides on the 3’ side of the mutation. The reverse oligo (5’ GCTGCAGGTAGCCCACATAGTAAG 3’) was designed in the other direction so that the 5’ end annealed back-to-back with the forward oligo. Oligonucleotides were purchased via the Eurofins Genomics Website (Luxembourg, EU). PCR was performed using Phusion HF PCR Master Mix (ThermoFisher Scientific). Cycling conditions were performed according to manufacturer instructions, except elongation which was done at 1 min/kb. PCR products sizes were verified by running samples on 1% agarose gel. Correctly amplified products were treated by Kinase-Ligase-Dpn1 (KLD) enzyme mix (New England Biolabs, Ipswich, MA, USA) for 30 min at room temperature. Samples were then transformed into XL10-Gold Ultracompetent cells (Agilent Technologies Santa Clara, CA, US) by heat shock and cells were plated on LB-Agar plate with Carbenicillin antibiotic (50 μL/mL; ThermoFisher Scientific). Plasmid DNA was extracted from antibiotic resistant colonies using a HiPure Miniprep kit (ThermoFisher Scientific). DNA samples were sent to Eurofins Genomics sequencing platform using the following primers:

- Ef1a For: 5’ TCAAGCCTCAGACAGTGGTTC 3’
- Spike seq primer 1(p800nt): 5’ CTCTGCTCTGGAACCCCTGGT 3’
- Spike seq primer 2(p1500nt): 5’ GAGTGTCTGTGATCACCCCT 3’
- Spike seq primer 3(p2500nt): 5’ CGGCTTCAATTTCAGCCAGATT 3’
- Spike seq primer 4(p2800nt): 5’ CAACAGCGCCATCGGCAAGA 3’
- rCD2 Rev: 5’ AACTTGCACCGCATATGCAT 3’

### T-cell activation assays

Prior to assays, T-cells were rested in R5 (as for R10 with 5% FBS) for 24 h before setting-up any activation assay to help reduce spontaneous activation. Typically, 3 ×10^4^ T-cells and 6 ×10^4^ target cells were used per well in R5 in a 96U well and incubated overnight, with supernatants collected for an ELISA (MIP-1β), which was performed according to the manufacturer’s instructions (R&D Systems) using half area ELISA plates. For TNFα-processing inhibitor (TAPI)-0 (Merck Group) assays, T-cells and target cells were co-incubated for 4 h with 30 μM TAPI-0 (Sigma, UK), antibody directed against TNF (clone cA2, Miltenyi Biotec) and CD107a antibody (clone H4A3, BD Biosciences) to detect activation-induced degranulation of cytotoxic T-cells. Following co-incubation, cells were washed and stained with Fixable Live/Dead Violet Dye VIVID and antibodies against appropriate cell surface markers as above.

### Peptide-MHC tetramer production and staining

Peptide-HLA-A*02:01 tetramers were produced and used to stain cells as previously described^52^. Briefly tetramers were assembled from monomers via the step-wise addition of PE-conjugated streptavidin (ThermoFisher Scientific) at a peptide-MHC:streptavidin molar ratio of 4:1. Protease Inhibitor (Merck Group) and PBS were added to give a final peptide-MHC tetramer concentration of 0.1 mg/ml (with regard to the peptide-MHC component). Following assembly, 0.5 μg of tetramer (with respect to the peptide-MHC component) was used to stain 5 ×10^4^ of a T cell clone and 4 ×10^6^ PBMCs. T-cell clones were stained from culture, COVID-19 patient PBMCs from cryopreserved samples and vaccinee PBMCs from freshly drawn blood. An optimised tetramer staining protocol involving pre-incubation with 50 nM protein kinase inhibitor, Dasatinib (Axon Medchem, Reston, VA, USA) and unconjugated anti-PE antibody (PE001, BioLegend) was used as described previously^53-55^. Following tetramer staining, T-cells were stained with LIVE/DEAD™ fixable violet dead stain (ThermoFisher Scientific), anti-CD3 and anti-CD8 antibody and analysed using flow cytometry as above. PBMCs were also stained with anti-CD14 (M5E2, BioLegend) and anti-CD19 (HIB19, BioLegend) pacific blue conjugated antibodies to create a ‘dump’ channel for dead cells.

### HLA A2 peptide binding assay

T2 cells were plated in a 96U-well plate (2 ×10^4^ per well) in serum-free media. Wuhan-YLQPRTFLL and P272L variant-YLQ**<u>L</u>**RTFLL Peptides (Peptide Protein Research, Fareham, UK) were added at 100 μM concentrations; alongside a negative control peptide HPVGEADYFEY from EBV that binds HLA B*35:01. The percentage volume of DMSO was matched in each condition. Cells were then incubated overnight at room temperature. The following day, cells were stained with anti-HLA A2 antibody (clone BB7.2, BioLegend) and incubated at 37°C for 1 h. Cells were then washed in PBS and stained with LIVE/DEAD™ fixable violet dead stain (ThermoFisher Scientific) and analysed using flow cytometry.

### TCR sequencing

TCR sequencing was performed as previously described^56^. RNA extraction was carried out using the RNEasy Micro kit (Qiagen, Hilden, Germany). cDNA was synthesized using the 5′/3′ SMARTer kit (Takara Bio, Paris, France). The SMARTer approach used a Murine Moloney Leukaemia Virus (MMLV) reverse transcriptase, a 3′ oligo-dT primer and a 5′ oligonucleotide to generate cDNA templates flanked by a known, universal anchor sequence at the 5′. PCR was then set up using a single primer pair consisting of TRAC or TRBC-specific reverse primer and an anchor-specific forward primer (Takara Bio, Paris, France). All samples were used for the following PCR reaction: 2.5 μL template cDNA, 0. 5 μL High Fidelity Phusion Taq polymerase, 10 μL 5X Phusion buffer, 0.5 μL DMSO, 1 μL dNTP Mix (10 mM each) (all from ThermoFisher Scientific, UK), 1 μL of TRAC or TRBC-specific primer (10 μM), 5 μL of 10X anchor-specific UPM primer (Takara Bio, Paris, France), and nuclease-free water for a final reaction volume of 50 μL (cycling conditions: 5 min at 94°C, 30 cycles of 30 s at 94°C, 30 s at 63°C for alpha chains or 30 s at 66°C for beta chains, 120 s at 72°C). Subsequently, 5 μL of the first PCR products were taken out to set up a nested PCR as above, using 1 μL of a nested primer pair (10 μM). All nested primers were flanked with Illumina indexes (cycling conditions: 5 min at 94°C, 30 cycles of 30 s at 94°C, 30 s at 62°C, 120 s at 72°C, and a final 10 min at 72°C). The final PCR products were loaded on a 1% agarose gel and purified with the Monarch® gel extraction kit (New England Biolabs). Purified products were sequenced on an Illumina MiSeq instrument using the MiSeq v2 reagent kit (Illumina, Cambridge, UK). Sequence analysis was performed using MiXCR software (v3.0.7)^57^. Public TCR clonotypes were identified using the VDJdb database^58^.

### Crystal structure determination

HLA A*02:01-YLQPRTFLL and HLA A*02:01-YLQLRTFLL protein crystals were grown at 18°C by sitting drop vapour diffusion as described^41^. 200 nL of protein (10 mg/ml) in crystallization buffer (10 mM Tris pH 8.1 and 10 mM NaCl) was added to 200 nL of reservoir solution. HLA A*02:01-YLQPRTFLL crystals grew in 0.2 M sodium nitrate, 0.1 M Bis Tris propane pH7.5, and 20 % w/v PEG 3350^59^. HLA A*02:01-YLQLRTFLL crystals grew in 0.2 M sodium iodide, 0.1 M Bis Tris propane pH7.5, and 20 % w/v PEG 3350^59^. Crystallization screens were conducted using an Art-Robbins Gryphon dispensing robot (Alpha Biotech Ltd, Killearn, Scotland UK). X-ray diffraction data were collected at 100 K at the Diamond Light Source (DLS), Oxfordshire, UK at a wavelength of ∼0.91808 Å using a PILATUS 6M pixel detector at beamline I04-1. Reflection intensities were estimated using Xia2, XDS, Dials, and Autoproc. The data were analysed with the CCP4 package^60^. Structures were solved with molecular replacement using PHASER^61^. Sequences were adjusted with COOT^62^ and the models refined with REFMAC5^63^. Graphical representations were prepared with the PYMOL molecular graphics system, version 2.3.4. The reflection data and final coordinates were deposited in the PDB database www.rcsb.org (HLA A*02:01-YLQPRTFLL PDB: 7P3D and HLA A*02:01-YLQ**<u>L</u>**RTFLL PDB: 7P3E).

### Data display

Unless stated otherwise, all data were displayed using GraphPad Prism software. TCR V-J usage plots were generated using VDJ tools^64^.

## Supporting information

PBD validation report for P272L peptide bound to HLA A*0201

PBD validation report for Wuhan peptide bound to HLA A*0201

GOG consortium

B.1.177 sequences

B.1.1.7 sequences

## Data Availability

All data generated or analysed during this study are included in this published article (and its supplementary information files). Raw data are available from the corresponding authors on reasonable request.

## ACKNOWLEDGEMENTS

This work was generously funded by Continuum Life Sciences (CLS) to aid the global COVID-19 effort. CLS provided salary for GD and CR. AKS is a Wellcome Investigator (220295/Z/20/Z) and we acknowledge salary support from Wellcome. EB received support from a Wellcome Trust Institutional Strategic Support Fund Fellowship. MSH was part funded by Enara Bio. We are grateful of funding from the Welsh Government through their Senior Research Leaders Scheme to AKS. LCJ is a Clinical Investigator at Cwm Taf Morgannwg University Health Board (CTMUHB). CTMUHB sponsored and supported the research and provided salary support for LCJ. TRC acknowledges support from the MRC, which funded the computational resources used by the project (grant reference MR/L015080/1), as well as specific funding from Welsh Government, which provided funds for the sequencing and analysis of Welsh samples used in this study, via Genomics Partnership Wales. UK sequencing has been supported by the COVID-19 Genomics UK Consortium (COG-UK), as well as using genomics data that has partly been funded/generated as part of COG-UK. COG-UK is supported by funding from the Medical Research Council (MRC) part of UK Research & Innovation (UKRI), the National Institute of Health Research (NIHR) and Genome Research Limited, operating as the Wellcome Sanger Institute. JAS is funded by the MRC (MR/T030062/1) and TW is funded by the Wellcome Trust (215800/Z/19/Z). The authors gratefully acknowledge the contributors to GISAID whose sequence data enabled an examination of the global spread of P272L (full list of sequences used and acknowledgements in supplementary information). The authors acknowledge Diamond Light Source for time on beamline I04-1 under proposal mx29502.

## AUTHOR CONTRIBUTIONS

GD, LCJ and AKS conceived the study. LCJ acquired the donor samples and clinical data. GD, CR, MSH, AW, BSz, EB, TW, JS, TM, KT, LRT, AF, PG, BSp, PJR, TC and AKS undertook experiments and/or analysed the data. GD, CR, TC and AKS wrote the manuscript.

---

**Supplementary Table 1:** A breakdown of numbers of sequenced cases possessing P272L in public databases, as determined by outbreak.info, downloaded on the 13^th^ of July 2021. The number of sequenced cases in public databases possessing P272L is broken down by PANGO lineage. B.1.177 and its sublineages are displayed below the other lineages where the mutation is found to simplify reading of the table. Lineages that have WHO variant designations are highlighted in dark grey (variants of concern) and mid grey (variants of interest) and light grey (alerts for further monitoring).

| PANGO Lineage | Number of sequenced cases from lineage in public databases | Number of cases with P272L in lineage | Proportion of cases with P272L in lineage | proportion ci lower | proportion ci upper | Percentage of cases in lineage |
| --- | --- | --- | --- | --- | --- | --- |
| B.1.1.7 | 982883 | 258 | 0.00026 | 0.00023 | 0.0003 | < 0.5% |
| B.1 | 83858 | 53 | 0.00063 | 0.00048 | 0.00082 | < 0.5% |
| B.1.160 | 25890 | 21 | 0.00081 | 0.00052 | 0.00122 | < 0.5% |
| B.1.429 | 36789 | 13 | 0.00035 | 0.0002 | 0.00059 | < 0.5% |
| B.1.351 | 27944 | 9 | 0.00032 | 0.00016 | 0.00059 | < 0.5% |
| B.1.404 | 634 | 7 | 0.01104 | 0.00495 | 0.02155 | 1% |
| B.1.1.263 | 626 | 6 | 0.00958 | 0.00401 | 0.01965 | 1% |
| B.1.1 | 47507 | 5 | 0.00011 | 4E-05 | 0.00023 | < 0.5% |
| B.1.526 | 47991 | 5 | 0.0001 | 4E-05 | 0.00023 | < 0.5% |
| B.1.1.214 | 17154 | 3 | 0.00017 | 4.9E-05 | 0.00047 | < 0.5% |
| B.1.1.63 | 2473 | 3 | 0.00121 | 0.00034 | 0.00323 | < 0.5% |
| B.1.2 | 95833 | 3 | 3.1E-05 | 8.8E-06 | 8.4E-05 | < 0.5% |
| B.1.524 | 585 | 2 | 0.00342 | 0.00071 | 0.01092 | < 0.5% |
| AD.2 | 4474 | 1 | 0.00022 | 2.4E-05 | 0.00104 | < 0.5% |
| B.1.1.153 | 852 | 1 | 0.00117 | 0.00013 | 0.00547 | < 0.5% |
| B.1.1.161 | 278 | 1 | 0.0036 | 0.00039 | 0.01669 | < 0.5% |
| B.1.1.174 | 250 | 1 | 0.004 | 0.00043 | 0.01854 | < 0.5% |
| B.1.1.222 | 3535 | 1 | 0.00028 | 3.1E-05 | 0.00132 | < 0.5% |
| B.1.1.39 | 1737 | 1 | 0.00058 | 6.2E-05 | 0.00269 | < 0.5% |
| B.1.1.519 | 21545 | 1 | 4.6E-05 | 5E-06 | 0.00022 | < 0.5% |
| B.1.1.523 | 466 | 1 | 0.00215 | 0.00023 | 0.00999 | < 0.5% |
| B.1.1.54 | 308 | 1 | 0.00325 | 0.00035 | 0.01507 | < 0.5% |
| B.1.160.28 | 330 | 1 | 0.00303 | 0.00033 | 0.01408 | < 0.5% |
| B.1.160.32 | 211 | 1 | 0.00474 | 0.00051 | 0.02193 | < 0.5% |
| B.1.170 | 105 | 1 | 0.00952 | 0.00103 | 0.04364 | 1% |
| B.1.214.3 | 110 | 1 | 0.00909 | 0.00098 | 0.0417 | 1% |
| B.1.221 | 13251 | 1 | 7.5E-05 | 8.1E-06 | 0.00035 | < 0.5% |
| B.1.222 | 579 | 1 | 0.00173 | 0.00019 | 0.00804 | < 0.5% |
| B.1.351.2 | 1958 | 1 | 0.00051 | 5.5E-05 | 0.00238 | < 0.5% |
| B.1.396 | 1274 | 1 | 0.00078 | 8.5E-05 | 0.00366 | < 0.5% |
| B.1.443 | 142 | 1 | 0.00704 | 0.00076 | 0.03244 | 1% |
| B.1.480 | 356 | 1 | 0.00281 | 0.0003 | 0.01305 | < 0.5% |
| B.1.596 | 9719 | 1 | 0.0001 | 1.1E-05 | 0.00048 | < 0.5% |
| B.1.617.2 | 155668 | 1 | 6.4E-06 | 6.9E-07 | 3E-05 | < 0.5% |
| C.36.3 | 1352 | 1 | 0.00074 | 8E-05 | 0.00345 | < 0.5% |
| P.1 | 50616 | 1 | 2E-05 | 2.1E-06 | 9.2E-05 | < 0.5% |
| B.1.177 | 72357 | 4704 | 0.06501 | 0.06323 | 0.06682 | 7% |
| B.1.177.10 | 1146 | 33 | 0.0288 | 0.02027 | 0.0397 | 3% |
| B.1.177.12 | 6593 | 7 | 0.00106 | 0.00048 | 0.00208 | < 0.5% |
| B.1.177.15 | 1496 | 2 | 0.00134 | 0.00028 | 0.00428 | < 0.5% |
| B.1.177.16 | 1452 | 14 | 0.00964 | 0.00554 | 0.01569 | 1% |
| B.1.177.17 | 1189 | 1 | 0.00084 | 9.1E-05 | 0.00392 | < 0.5% |
| B.1.177.21 | 13047 | 1 | 7.7E-05 | 8.3E-06 | 0.00036 | < 0.5% |
| B.1.177.23 | 292 | 1 | 0.00342 | 0.00037 | 0.01589 | < 0.5% |
| B.1.177.31 | 78 | 3 | 0.03846 | 0.01095 | 0.09908 | 4% |
| B.1.177.33 | 373 | 7 | 0.01877 | 0.00843 | 0.03649 | 2% |
| B.1.177.35 | 1607 | 1 | 0.00062 | 6.7E-05 | 0.0029 | < 0.5% |
| B.1.177.40 | 200 | 1 | 0.005 | 0.00054 | 0.02313 | 1% |
| B.1.177.44 | 1793 | 5 | 0.00279 | 0.00106 | 0.0061 | < 0.5% |
| B.1.177.50 | 522 | 2 | 0.00383 | 0.0008 | 0.01223 | < 0.5% |
| B.1.177.54 | 1602 | 1 | 0.00062 | 6.7E-05 | 0.00291 | < 0.5% |
| B.1.177.69 | 556 | 1 | 0.0018 | 0.00019 | 0.00838 | < 0.5% |
| B.1.177.7 | 5949 | 4 | 0.00067 | 0.00023 | 0.0016 | < 0.5% |
| B.1.177.72 | 279 | 1 | 0.00358 | 0.00039 | 0.01663 | < 0.5% |
| B.1.177.75 | 784 | 1 | 0.00128 | 0.00014 | 0.00595 | < 0.5% |
| B.1.177.81 | 4368 | 1 | 0.00023 | 2.5E-05 | 0.00107 | < 0.5% |
| B.1.177.82 | 1908 | 1885 | 0.98795 | 0.98228 | 0.99213 | 99% |
| B.1.177.83 | 543 | 542 | 0.99816 | 0.99142 | 0.9998 | 100% |
| B.1.177.84 | 149 | 136 | 0.91275 | 0.85947 | 0.95021 | 91% |
| B.1.177.86 | 5319 | 4 | 0.00075 | 0.00025 | 0.00179 | < 0.5% |
| B.1.177.87 | 1302 | 1257 | 0.96544 | 0.95446 | 0.97435 | 97% |
| B.1.177.88 | 70 | 68 | 0.97143 | 0.9115 | 0.99402 | 97% |
| B.1.177.89 | 44 | 43 | 0.97727 | 0.89866 | 0.99754 | 98% |

**Supplementary Table 2.** Data collection and refinement statistics for Human MHC I, A*02 allele, presenting Wuhan (YLQPRTFLL) and escape mutant (YLQ**<u>L</u>**RTFLL) SARS-Cov-2 Spike protein epitope

| **PDB Entry** | **7P3D** | **7P3E** |
| --- | --- | --- |
| **Data Collection\*** |  |  |
| Diamond Beamline | I04-1 | I04-1 |
| Date | 2021-06-28 | 2021-06-28 |
| Wavelength | 0.91808 | 0.91808 |
| **Crystal Data (figures in brackets refer to outer resolution shell)** |  |  |
| Crystallisation Conditions | 0.2 M Na Nitrate, 0.1 M Bis-Tris propane, 20% PEG3350, pH 7.5 | 0.2 M NaI, 0.1 M Bis-Tris propane, 20% PEG3350, pH 7.5 |
| *a,b,c* (Å) | 56.63, 79.42, 57.89 | 85.80, 85.80, 217.56 |
| $\alpha,\beta,\gamma$ (°) | 90.0, 116.44, 90.0 | 90.0, 90.0, 120.0 |
| Space group | P 1 2<sub>1</sub> 1 | P 3<sub>2</sub> 1 2 |
| Resolution (Å) | 1.67 – 51.89 | 2.00 – 74.31 |
| Outer shell | 1.67 – 1.70 | 2.00 - 2.05 |
| *R*-merge (%) | 16.2 (141.9) | 8.6 (109.4) |
| *R*-pim (%) | 9.5 (83.9) | 3.7 (47.0) |
| *R*-meas (%) | 18.8 (165.6) | 9.0 (114.5) |
| CC1/2 | 0.990 (0.364) | 0.998 (0.779) |
| *I* / $\sigma$(*I*) | 9.5 (0.9) | 15.1 (2.3) |
| Completeness (%) | 99.9 (100.0) | 99.7 (96.3) |
| Multiplicity | 3.9 (3.9) | 11.5 (11.8) |
| Total Measurements | 208,201 (10,359) | 720,769 (52,255) |
| Unique Reflections | 53,176 (2,648) | 62,446 (4,415) |
| Wilson B-factor(Å<sup>2</sup>) | 13.8 | 38.8 |
| **Refinement Statistics** |  |  |
| Refined atoms | 3,651 | 6,517 |
| Protein atoms | 3,173 | 6,348 |
| Non-protein atoms | 29 | 38 |
| Water molecules | 449 | 131 |
| R-work reflections | 50,389 | 59,258 |
| R-free reflections | 2,697 | 3,051 |
| R-work/R-free (%) | 18.9 / 23.1 | 21.4 / 24.8 |
| **rms deviations (target in brackets)** |  |  |
| Bond lengths (Å) | 0.012 (0.013) | 0.013 (0.013) |
| Bond Angles (°) | 1.586 (1.663) | 1.464 (1.657) |
| <sup>1</sup>Coordinate error | 0.103 | 0.147 |
| Mean B value (Å<sup>2</sup>) | 20.1 | 47.0 |
| **Ramachandran Statistics (PDB Validation)** |  |  |
| Favoured/allowed/Outliers | 381 / 8 / 0 | 741 / 16 / 2 |
| % | 98.0 / 2.0 / 0.0 | 98.0 / 2.0 / 0.0 |

\* One crystal was used for determining each structure.
<sup>1</sup> Coordinate Estimated Standard Uncertainty in (Å), calculated based on maximum likelihood statistics.

**Supplementary Figure 1.**
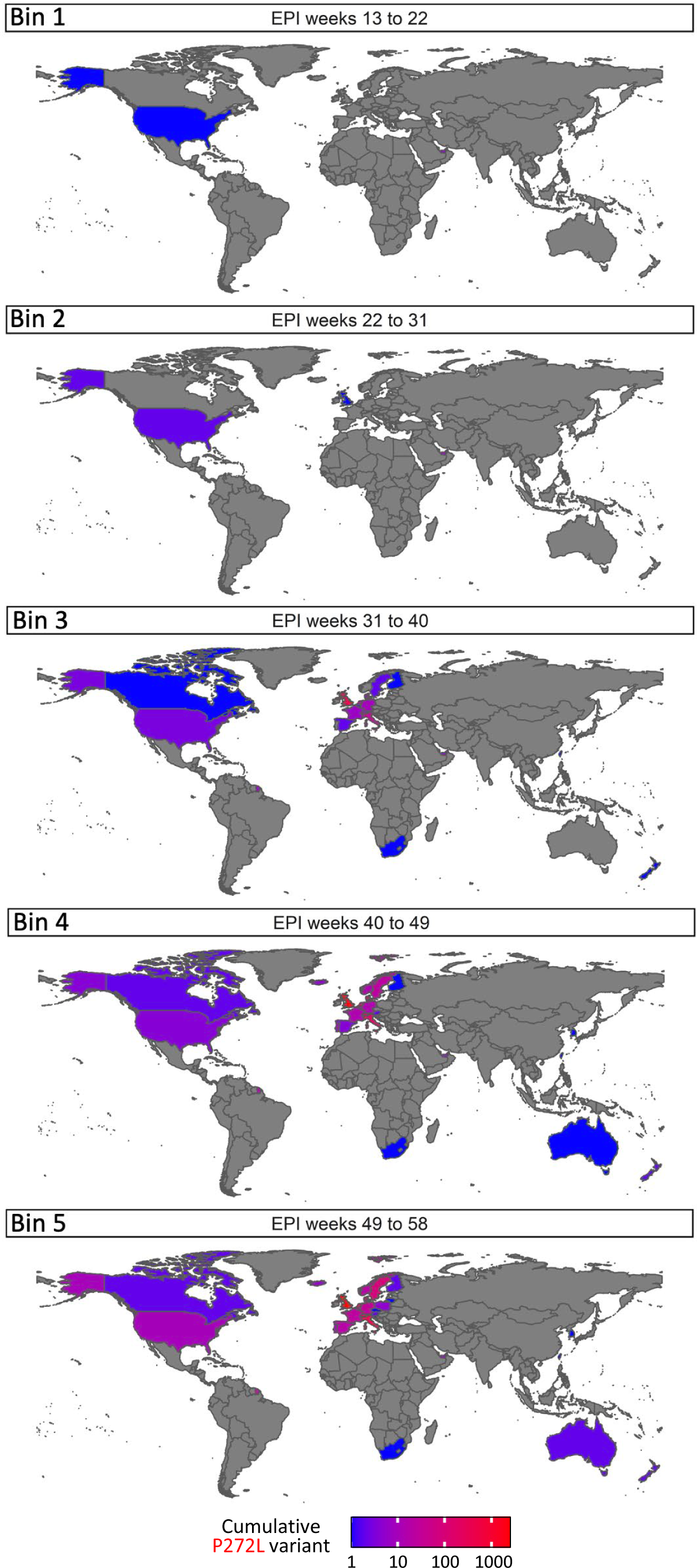
Worldwide SARS-CoV-2 viral dynamics of P272L spike mutation. Total number of P272L variants by nation, binned into periods of 10 epidemiological (EPI) weeks, from EPI week 13 (beginning 22^nd^ March 2020), the first time a sequence with P272L was observed in sequence data; up to and including January 31^st^ 2021 (EPI week 58).

**Supplementary Figure 2.**
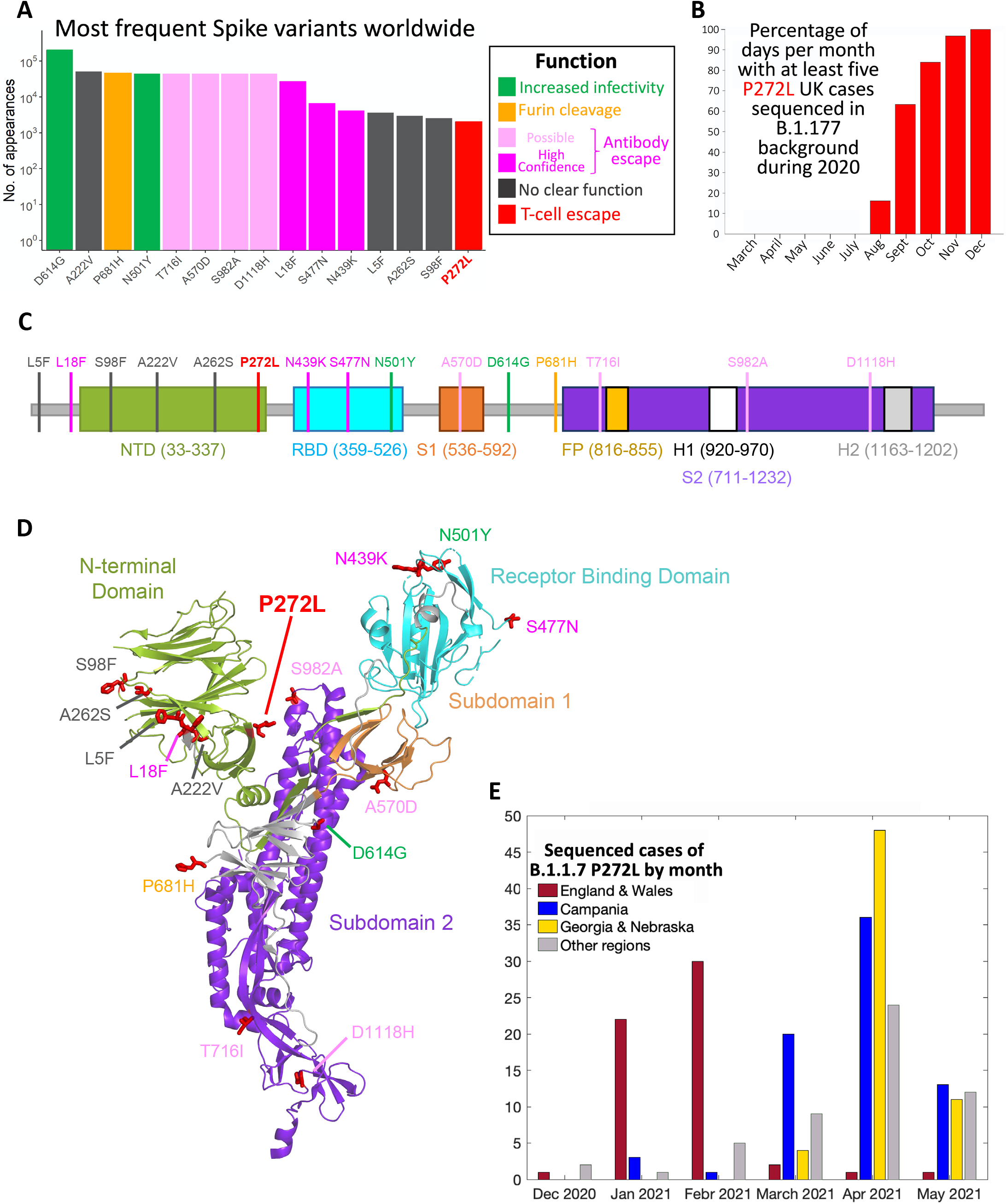
**(A**) Top 15 most frequently observed variants observed in worldwide spike glycoprotein sequence data in all lineages. Those thought to be associated with enhanced viral transmission are shown in green and those listed as potentially escaping from antibody mediated-mediated immunity are shown in light pink (possible) and dark pink (high confidence) as assigned at http://sars2.cvr.gla.ac.uk/cog-uk/#shiny-tab-immunology. This colour-coding of substitutions is used throughout the figure. Variants of unknown function in grey. N501Y also escapes from antibodies **(B**) Fraction of days per month where five or more P272L variants were sequenced in B.1.177 background. **(C**&**D**) Mapping of mutants shown in A onto Spike sequence map and prefusion structure (PDB 6VXX; Walls et al. 2020. *Cell*, **181**, 281-291). **(E**) Localised emergence of P272L in the B.1.1.7 lineage prior to dominance of B.1.617.2 (Delta) variant.

**Supplementary Figure 3.**
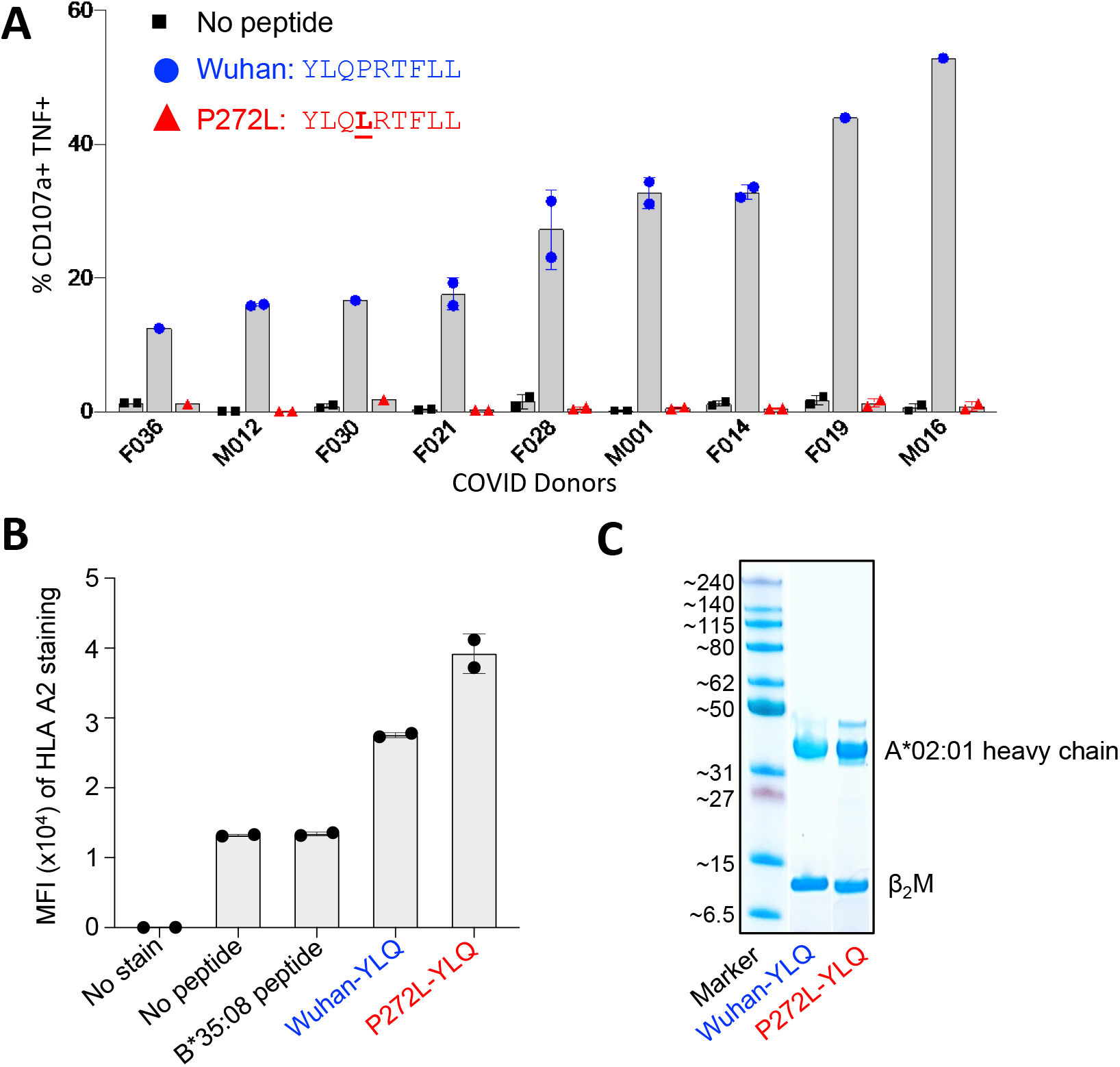
Wuhan-YLQPRTFLL T-cell lines from SARS-CoV-2 convalescent patients do not activate with P272L-YLQ<u>L</u>RTFLL variant peptide. **(A**) PBMCs from nine SARS-CoV-2 CP were enriched with Wuhan-YLQPRTFLL tetramers then expanded with allogeneic PBMC feeders and PHA. The lines were used in a 4h T107a assay with 10^−6^ M Wuhan-YLQPRTFLL or P272L-YLQ**<u>L</u>**RTFLL peptides. **(B**) Both Wuhan and P272L peptides bind to HLA A2. HLA A2 APC staining data from a T2 binding assay using Wuhan-YLQPRTFLL and P272L-YLQ**<u>L</u>**RTFLL exogenous peptides. HLA B*35:08 binding HPVGEADYFEY peptide used as an irrelevant control. Error bars depict SD of duplicates. **(C**) Refolded Wuhan-YLQPRTFLL and P272L-YLQ**<u>L</u>**RTFLL HLA monomers.

**Supplementary Figure 4.**
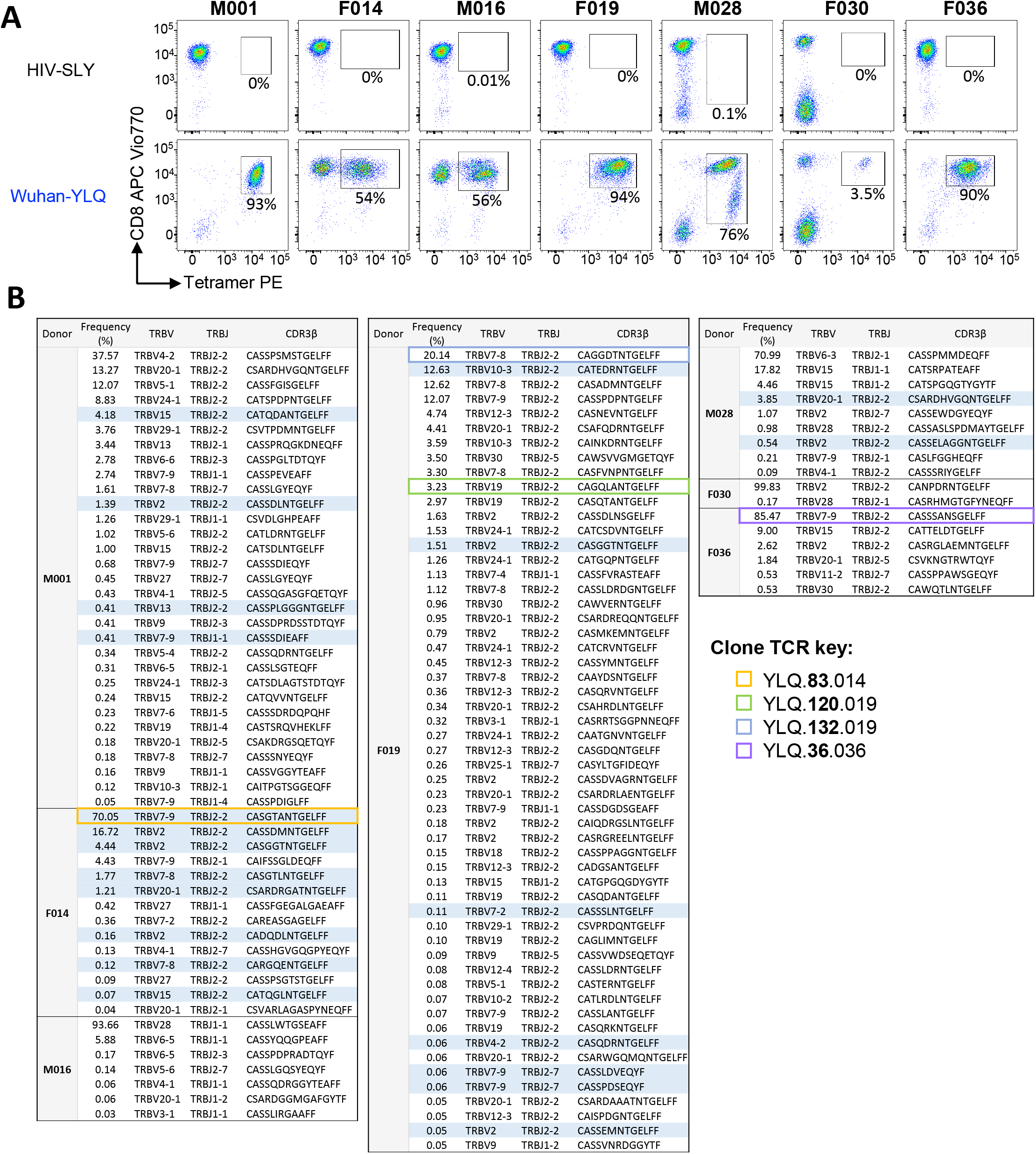
TCR usage and CDR3s from Wuhan-YLQPRTFLL tetramer sorted T-cells from lines generated from seven SARS-CoV-2 patients. **(A**) Wuhan-YLQPRTFLL tetramer sorted T-cell lines for TCR sequencing shown in **Figure 3C. (B**) TCR sequences of the Wuhan-YLQPRTFLL specific T-cells. Public TCR chains are highlighted in blue. TCR chains of the T-cell clones used in the study are indicated according to the key.

**Supplementary Figure 5.**
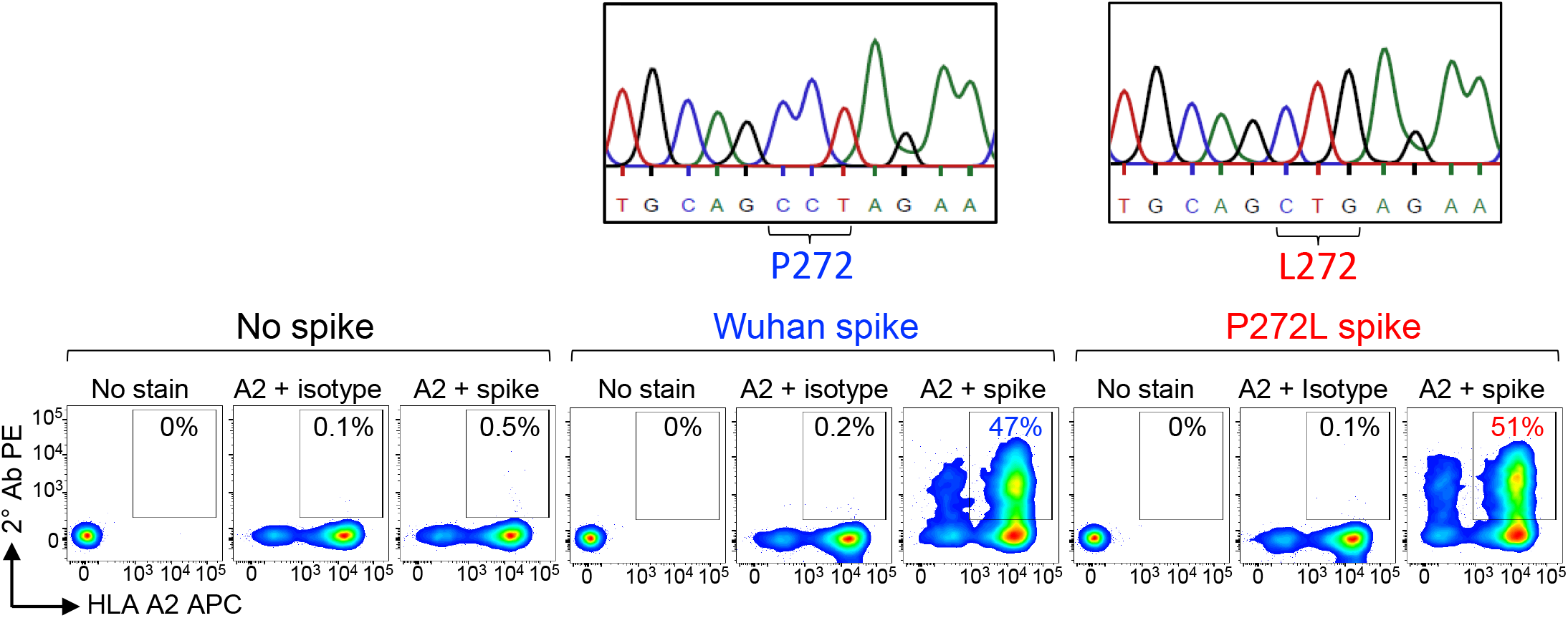
Confirmation of HLA A2 and spike expression of transduced A549 cells. HLA A2 (A2) and spike protein expression of the Wuhan and P272L spike transduced A549-A2. HLA A2 staining with BB7.2 Ab clone. Unconjugated isotype and anti-spike antibodies used in conjunction with a PE conjugated secondary antibody on Y axis.

**Supplementary Figure 6.**
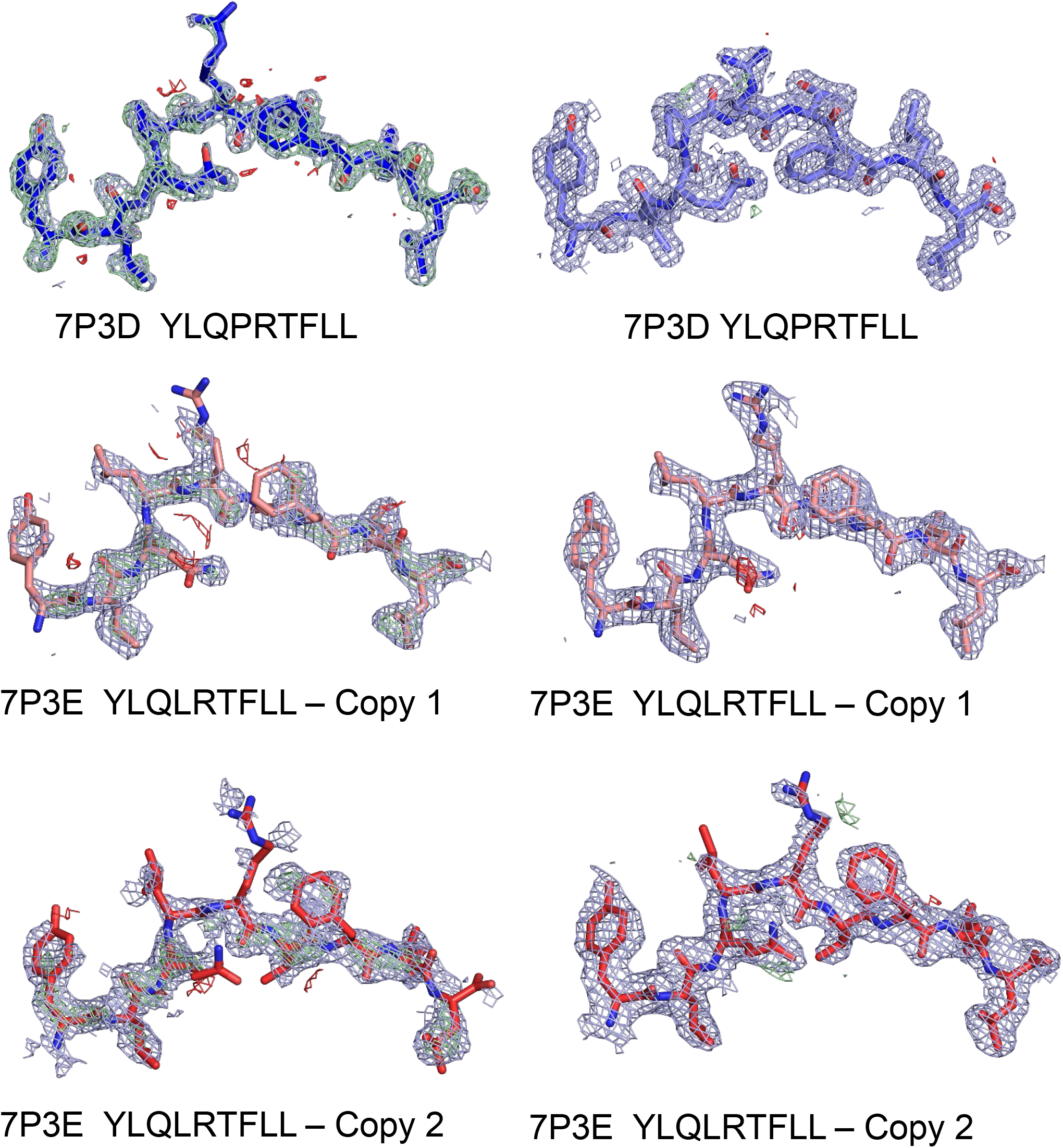
Electron density maps around the peptides in the MHC structures. The left column shows the final coordinates of the peptide in an omit map obtained after solving each structure with PHASER using a model not including the peptide. The right column shows the electron density in the final map obtained after convergence of refinement in REFMAC. The density is displayed in light blue from the observed map, in green contours from the positive difference map, and in red for the negative contours from the difference map.

